# Reduced response to SARS-CoV-2 vaccination is associated with impaired immunoglobulin class switch recombination in SLE patients

**DOI:** 10.1101/2024.08.14.24310924

**Authors:** Guillem Montamat, Claire E. Meehan, Hannah F. Bradford, Reşit Yıldırım, Francisca Guimarães, Marina Johnson, David Goldblatt, David A. Isenberg, Claudia Mauri

**Affiliations:** Division of Infection and Immunity and Institute of Immunity and Transplantation, Royal Free Hospital, University College London, UK; Centre for Rheumatology, Division of Medicine, University College London Hospital, London, UK; Great Ormond Street Institute of Child Health Biomedical Research Centre, University College London, London, UK

## Abstract

**Objective:** Systemic Lupus Erythematosus (SLE) patients exhibit B-cell abnormalities. Although there are concerns about reduced antibody responses to SARS-CoV-2 vaccines, detailed data on B-cell-specific responses in SLE remain scarce. Understanding the responsiveness to novel vaccine-antigens, and boosters number, is important to avoid unnecessary prolonged isolation of immunocompromised individuals. We assessed humoral and antigen-specific B-cell subset responses, including changes in isotype switching, prior to and after several doses of SARS-CoV-2 vaccines.

**Methods:** Blood samples were obtained prior to and after SARS-CoV-2 vaccination from cross-sectional and longitudinal cohorts of previously uninfected patients with SLE (n=93). Healthy participants receiving SARS-CoV-2 vaccines were recruited as controls (n=135). We measured serum antibody titres, their neutralizing capacity, and vaccine-specific memory B cells subsets.

**Results:** Impaired IgG, IgA, and neutralizing responses against the original and various SARS-CoV-2 variants were observed following two doses of vaccine in SLE patients. Follow-up booster doses increased humoral responses compared to baseline, but they remained lower, with poorer neutralisation capacity against most strains, compared to healthy individuals after three or more doses. Analysis of memory B-cells subsets in SLE patients revealed an increase of SARS-CoV-2-specific isotype unswitched IgM^+^ over SARS-CoV-2-specific isotype switched IgG^+^/IgA^+^memory B-cells compared to healthy individuals. Culturing healthy naive B-cells with high levels of IFNα, a hallmark of SLE pathogenesis, prevented B-cells from switching to IgG under IgG-polarizing conditions.

**Conclusion:** SLE patients’ protection against SARS-CoV-2 is overall impaired compared to healthy individuals and is associated with a class switch defect possibly due to chronic exposure of B-cells to IFNα.

**GRAPHICAL ABSTRACT:** 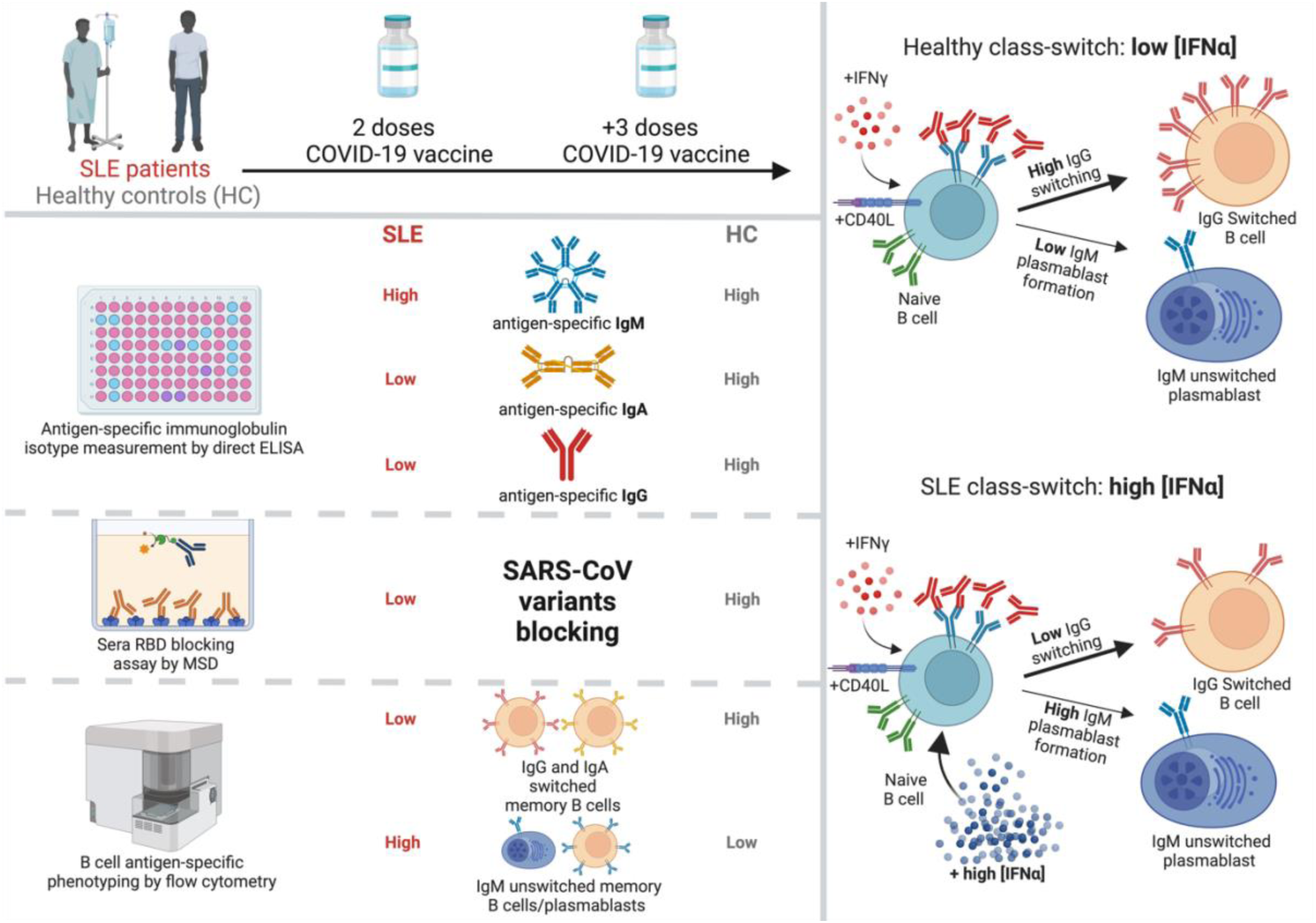

## INTRODUCTION

Systemic lupus erythematosus (SLE) is a chronic and clinically heterogeneous autoimmune disease, characterised by the presence of pathogenic autoantibodies. SLE patients present with profound B cell abnormalities (1), including expansions of pathogenic CD27^-^IgD^-^double-negative (DN) B cells, autoantibody-producing plasmablasts, and by reduced frequencies of anti-inflammatory regulatory B cells (Bregs) (2). B cell dysregulation is often associated to more severe disease and to high autoantibody titres.

During infection, early extrafollicular B cell reactions, resulting in IgM production (3), provide initial protection against pathogens, followed by the generation of long-lasting B cell immunity occurring through follicular activation including the germinal centre (GC) reaction (4). GC formation and the consequent development of somatically hypermutated memory B cells and plasma cells are pivotal in driving efficacious protective immunity not only following infection but also following vaccination. Follicular responses facilitate class switch recombination (CSR), leading to the production of more functional IgG and IgA antibodies (5). It has been previously reported that SARS-CoV-2 mRNA vaccination induces a persistent GC response that can lasts up to six months in the general population (6).

Comprehensive characterization of the antibody-mediated immune reaction in SLE patients, including both antibody quality and detailed B cell response, remains largely unexplored (7–12). To bridge this gap in knowledge, we compared the efficacy of SARS-CoV-2 vaccination in a cohort of SLE patients and healthy individuals by measuring serological and vaccine-specific B cell responses.

In agreement with previous findings (7), we show a reduced response to SARS-CoV-2 vaccination following the initial 2 doses. Compared to healthy controls, SLE patients exhibit lower receptor-binding domain (RBD)- or spike protein subunit 1 (S1)-specific IgG titres with weaker neutralizing capacity. We reiterate the importance of booster vaccinations for immunocompromised individuals in building sufficient functional antibody responses. However, we report that SLE patients present a CSR defect in vaccine-specific B cells compared to healthy individuals as shown by an accumulation of vaccine-specific IgM^+^unswitched memory B cells. We recapitulate the CSR defect by culturing healthy B cells with high levels of IFNα, a major contributor to B cell abnormalities in SLE, under IgG isotype switching polarising conditions. Taken together our results show that IFNα-induced defective CSR could underpin to sub-optimal immune response to vaccination in SLE patients.

## METHODS

### Study population

Peripheral blood samples for PBMC and serum isolation were collected from healthy donors and from SLE patients attending the University College London Hospital (UCLH) rheumatology outpatients’ clinic. Ethical approval was obtained from the UCLH Health Service Trust ethics committee, under REC reference no. 14/SC/1200. Healthy controls (n=135) and patients (n=93) were recruited following informed consent. Sample storage complied with requirements of the Data Protection Act 1998. Demographics, clinical features, routine laboratory testing and therapeutic regimen were collected retrospectively from electronical medical files. Dates of vaccination and history of SARS-CoV-2 infection were also recorded for healthy controls and SLE patients. Detailed exclusion criteria are described in online supplemental material.

### Quantification of RBD-specific, S1-specific, and total IgG, IgA and IgM titres

RBD- and S1-specific direct ELISA protocols were carried out as previously published 13 with modifications. Similarly, RBD-omicron-specific IgG was an adaption of this protocol using an anti-omicron IgG monoclonal antibody. RBD-omicron-specific IgA was determined using a commercial kit (RAS-T099, AcroBiosystems). To measure the total isotype (IgG, IgA, and IgM) immunoglobulin concentration in serum samples, a sandwich ELISA (88-50550-22 for IgG, 88-50600-88 for IgA, 88-50620-88 for IgM, Invitrogen) was performed according to the manufacturer’s instructions. Detailed protocols are described in the supplemental methods.

### Blocking assay

A V-PLEX SARS-CoV-2 panel 30 (ACE2) kit (K15635U, Meso Scale Diagnostics) was used according to the manufacturer’s instructions to measure the capacity of serum samples from SLE patients and healthy controls to block the binding of variants of SARS-CoV-2 to the ACE2 receptor. A dilution of 1:50 was used for all serum samples. Individuals with blocking capacity >50% for any tested SARS-CoV-2 strain were considered as neutralisers.

### *Ex vivo* B cell phenotyping

SARS-CoV-2 antigen-specific B-cells were identified using labelled RBD (793604, Biolegend) and S1 (792906, Biolegend) proteins with the addition of antibodies against phenotypic markers and BCR isotype-specific antibodies to measure class-switch status. Full antibody panels and protocols used for *ex vivo* B cell phenotyping is described in supplemental methods. Stained samples were acquired using a Cytek™ Aurora spectral cytometer, using SpectroFlo (v3.0.3) with automated unmixing. Data were analysed using FlowJo (TreeStar, v.10.8.1).

### IFNα CSR *in vitro* culture

Naive B cells were sorted from previously frozen PBMCs of healthy controls, using a BD FACSAria™ Fusion and BD FACSDiva (v9.4). Full details of cell sorting protocols are described in supplemental methods. Sorted naive B cells from healthy controls were cultured *in vitro* under IgG-polarising class-switching conditions, consisting of complete RPMI (10% heat-inactivated fetal calf serum, 1% penicillin/streptomycin), 1μg/ml anti-IgM (309-006-043, Jackson ImmunoResearch), 1μg/ml CD40L (ALX-522-110-C010, Enzo Life Sciences), and 50ng/ml recombinant human IFNγ (285-IF-100/CF, Bio-Techne), in a round-bottom 96-well plate at 37°C and 5% CO_2_ for 6 days. IgG polarising class-switch medium was refreshed on day 3 by gently centrifuging the cells (300g for 10min) and adding fresh media including the above-mentioned factors. Staining protocols and antibody panels used are described in supplemental methods. Stained samples were acquired using a Cytek™ Aurora spectral cytometer, using SpectroFlo (v3.0.3) with automated unmixing. Data were analysed using FlowJo (Treestar, v10.8.1).

### Statistical analysis

The normality of each dataset was determined using the Kolmogorov-Smirnov’s, Anderson-Darling’s, D’AgostinoPearson’s and Shapiro-Wilk’s tests, and the quantile-quantile (q-q) plot. For non-parametric datasets, Wilcoxon matched-pairs signed rank test was used to analyse paired data sets, Mann-Whitney for unpaired datasets, and Kruskal-Wallis test for multivariable datasets. For parametric datasets, a paired or unpaired t-test was used to analyse data and an ordinary one-way ANOVA was used for multivariable datasets. For parametric datasets where variance differed between groups, unpaired t-test with Welch’s correction was used. For non-parametric datasets where SD differed between groups, a log transformation to achieve normal distribution and tests for parametric data were used. Violin plots display the median, along with the 25th and 75th percentiles. Bar plots graphs are expressed as the mean +/- standard error of the mean. Graphing of data and analysis of statistical significance was performed in Prism (GraphPad v9) and R (version v4.2.2). Results were considered significant at \**P*<0.05, \*\**P*<0.01, \*\*\**P*<0.001, \*\*\*\**P*<0.0001.

## RESULTS

### Study cohort

Antibody responses to the SARS-CoV-2 vaccine were measured in a cross-sectional cohort of 93 SARS-CoV-2 naive SLE patients. 34 patients were recruited pre-pandemic (no vaccine), and 59 patients post vaccination after either 2 or 3^+^ doses of mRNA-based vaccine (Pfizer or Moderna, n=34); non-mRNA vector-based vaccine (Astrazeneca, n=7); or a combination of two vaccine types (n=15). 135 healthy volunteers were also recruited; 70 pre-pandemic (no vaccine), and 65 after either 2 or 3^+^ doses of mRNA-based vaccine (Pfizer or Moderna, n=57); non-mRNA vector-based vaccine (Astrazeneca, n=3); or a combination of two vaccine types (n=4). A small longitudinal cohort of 18 SLE patients and 22 healthy controls was also included. Further demographic details for the cross-sectional and longitudinal cohorts can be found in tables 1 and supplemental table 1, respectively.

**Table 1.**
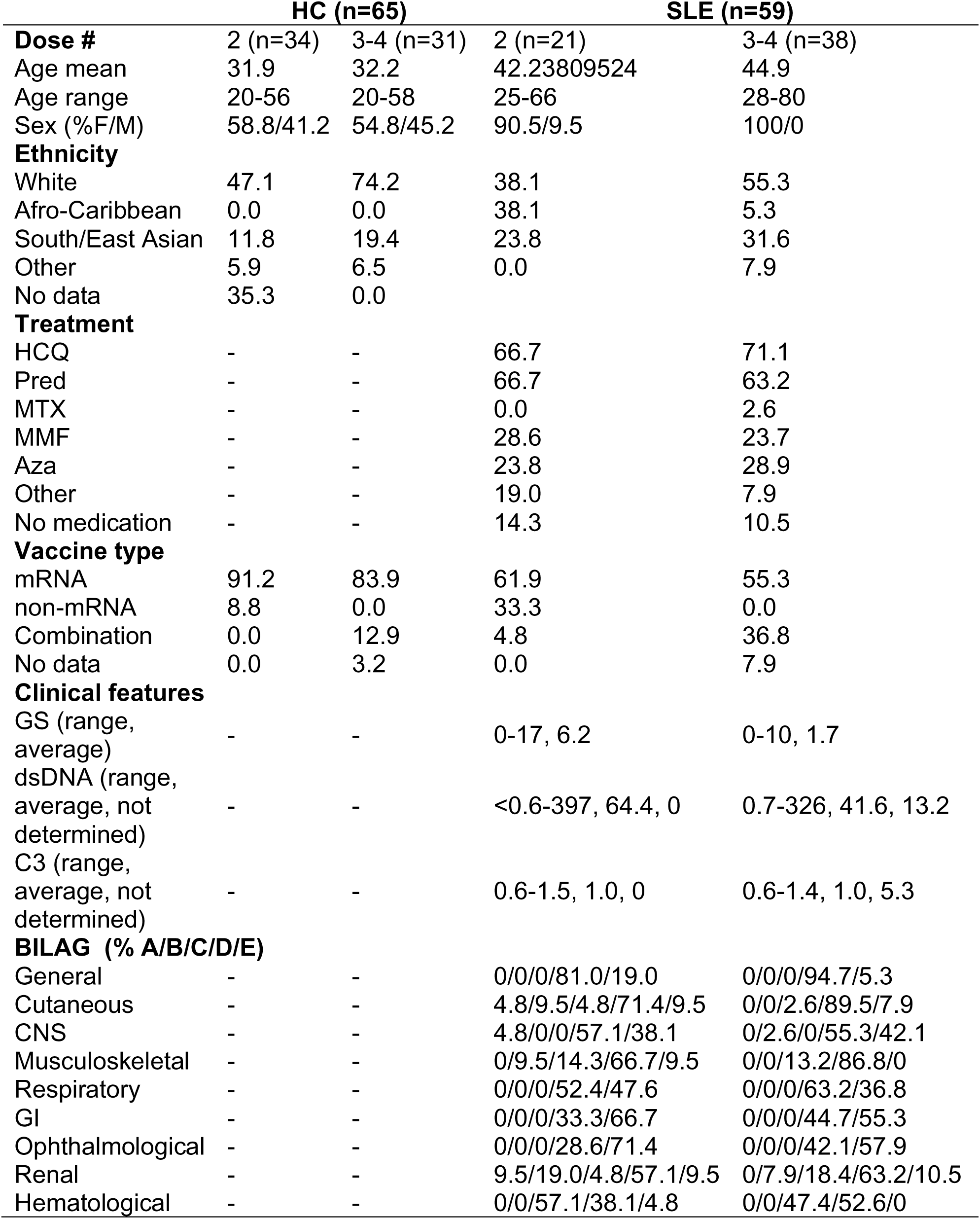
Demographics of cross-sectional cohort of SLE patients and healthy controls. HCQ = hydroxychloroquine, Pred = prednisolone, MTX = methotrexate, MMF = mycophenolate mofetil, Aza = azathioprine, GS = global score, dsDNA = double-stranded DNA, BILAG = British Isles Lupus Assessment Group, CNS = central nervous system, GI = gastrointestinal.

### SLE patients show reduced serum SARS-CoV-2 IgG antibody response compared to healthy individuals

The subunit 1 (S1) of the SARS-CoV-2 spike protein interacts with the ACE receptor on the host cell. This subunit includes the ACE receptor binding domain (RBD). Virus-specific (S1 and RBD) vaccine-derived IgG, IgA, and IgM levels (ng/ml) were assessed using direct ELISA. We report high degree of variability of vaccine responses for all antibody isotypes in both healthy controls and SLE patients (Figure 1A-C).

**Figure 1.**
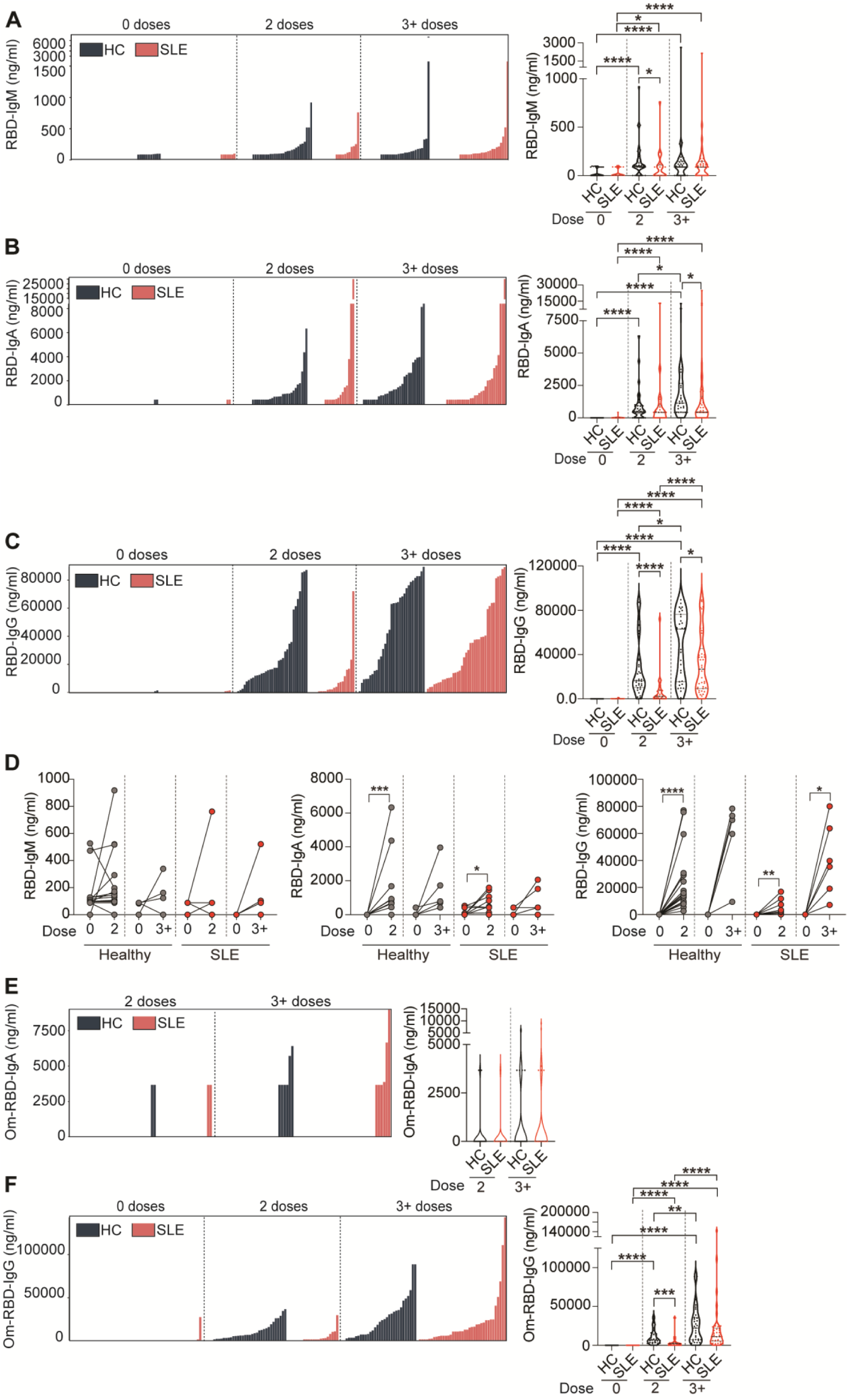
SLE patients have impaired humoral response after SARS-CoV-2 vaccination. **(A-C)** Individual data bar plots and cumulative data violin plots showing the titres of RBD-specific (A) IgM, (B) IgA, and (C) IgG antibodies in a cross-sectional cohort of SLE patients and healthy controls (HC), pre-vaccination and following 2 or 3^+^ doses of vaccine. **(D)** Graphs showing the titres of RBD-specific IgM, IgA and IgA antibodies in a longitudinal cohort of SLE patients and healthy controls (HC), between pre-vaccination (0 doses) and 2 or 3^+^ doses of vaccine. **(E,F)** Individual data bar plots and cumulative data violin plots showing the titres of Omicron (Om)-specific **(E)** IgA and **(F)** IgG antibodies in a cross-sectional cohort of SLE patients and healthy controls (HC), pre-vaccination and following 2 or 3^+^ doses of vaccine. \**P*<0.05, \*\**P*<0.01, \*\*\**P*<0.001, \*\*\*\**P*<0.0001, (A to F) by Mann-Whitney of selected group pairs or by Wilcoxon matched-pairs signed rank test. Violin plots display the median, along with the 25th and 75th percentiles.

Analysis of specific antibody isotypes showed that IgM responses against both RBD and S1 contributed minimally to the spike-reactivity in healthy and SLE cohort at any dose tested (Figure 1A, supplemental Figure 1A).

IgA-specific responses to RBD and S1 were impaired in SLE patients after 2 and 3^+^ doses as shown by reduced IgA median titers (IgA titre median after 2 doses: RBD HC=527.5 vs SLE=424.1; S1 HC=846.5 vs SLE=0; IgA titre median after 3^+^ doses: RBD HC=1145 vs SLE=437.1; S1 HC=1613 vs SLE=999.7) (Figure 1B, supplemental Figure 1B).

Although the titers of anti-RBD and anti-S1 IgG significantly increased following booster doses (3^+^doses) in the majority of SLE patients compared to the levels detected after the second dose, median levels remained significantly lower than those in healthy individuals (IgG titre median after 3^+^ doses: RBD HC=63k vs SLE=26.7k; S1 HC=113.7k vs SLE=53.5k) (Figure 1C, supplemental Figure 1C).

SLE patients are known to present hyper-globulinemia (14) and treatment-derived hypo-globulinemia (15), which might influence vaccine responses. To consider the “weight” that this defect in total antibodies might have on the vaccine-specific Ig response, we calculated the relative levels of antigen-specific IgM/A/G as a percentage of respective total Ig isotype. SLE patients present lower amount of total IgG and IgM and an increase in total IgA levels compared to healthy controls both before and after vaccination (supplemental Figure 2A). When calculated as percentage of the total respective Ig isotype, SLE patients continued to show lower RBD-IgG and IgM responses compared to healthy controls after 2 and 3^+^ doses, (supplemental Figure 2B,D). No significant differences were observed in the percentages of RBD-IgA/total IgA between SLE patients and healthy controls at any of the doses tested (supplemental Figure 2C).

Analysis of the longitudinal cohort confirmed that although the levels of RBD-specific IgA and IgG were significantly increased in SLE patients after vaccination, the medians of IgG and IgA titres were higher in healthy compared to SLE patients even after 3^+^dose (titre median after 3^+^ doses: RBD IgG HC=74.2k vs SLE=37.5k; IgA HC=704 vs SLE=212.1) (Figure 1D). No changes in IgM levels were recorded after vaccination in these samples (Figure 1D).

Several studies have indicated that immunosuppressant-drugs and or severity of disease are associated to weaker SARS-CoV-2 vaccination responses (9,11). No significant associations with antibody response across different treatments (DMARDs, including hydroxychloroquine (HCQ), or prednisolone) were observed at any of the doses tested (supplemental Figure 3A,B). We report that RBD-IgM titres were increased in patients with active disease (BILAG global scoring ≥5) compared to those with inactive disease (BILAG <5) after 3^+^ doses. No associations were found between disease activity and RBD-IgA nor RBD-IgG titres (supplemental Figure 3C).

SARS-CoV-2 vaccination targeting the ancestral strain has demonstrated limited efficacy against the Omicron variant in healthy individuals (16), however, the ability of SLE patients to respond to new variants remains largely unexplored. Omicron RBD-specific IgA (Om-RBD-IgA) titres were undetectable in the majority of SLE patients and healthy controls for any number of doses (Figure 1E). We confirm that IgG titres against the Omicron RBD were generally lower compared to the response against the original strain of SARS-CoV-2 after 2 doses in both healthy controls and SLE patients (Figure 1F). A moderate increase of IgG titres was seen in both groups after 3^+^ doses with healthy controls showing a higher median (Figure 1F, IgG titre median after 3^+^ doses: Om-RBD HC=22.2k vs SLE=12.2k).

### SLE patients exhibit impaired neutralising capacity against variants of concern

Next, we assessed the functional neutralisation capacity of circulating antibodies to block the RBD protein of various SARS-CoV-2 variants from binding to the ACE2 receptor by using an MSD assay in a cross-sectional cohort (SLE n=59, HC n=65) (demographic details in supplemental table 2). The analysis included the original strain (CoV.2) and several variants of concern (VOC), comprising Alpha (B.1.1.7), Beta (B.1.351), Delta, and a range of sub-Omicron lineages (BA.2, BA.4, BA.5, BA.2.12.1, BA.2.75). Reflecting the lower amount of anti-IgG production, viral neutralisation, and ratio of neutraliser individuals against all variants were very low in SLE patients after 2 doses (Figure 2A-D). Despite lower levels of antibodies detected in SLE patients compared to healthy controls after 3^+^ doses (represented here as alignment between serum IgG levels and respective neutralisation capacity in Figure 2B), the inhibition capacity for all but one (BA.4;BA.5) strain was not significantly different from healthy controls (Figure 2C). However, when taking into account the ratio of neutralising individuals, SLE patients showed a lower percentage of neutralisers for all strains after 3^+^ doses (HC=93.55% vs SLE=82.35% for CoV-2, HC=90.32% vs SLE=70.59% for B.1.1.7, HC=64.52% vs SLE=52.94% for B.1.351, HC=83.87% vs SLE=64.70% for Delta, HC=38.71% vs SLE=20.59% for BA.2, HC=51.61% vs SLE=29.41% for BA2.12.1 and HC=48.39% vs SLE=26.47% for BA.4/BA.5), but for the strain BA.2.75 (HC=12.9% vs SLE=11.76%) (Figure 2D). Thus, our results reinforce the need for booster doses to increase neutralizing capacity, mainly driven by IgG titres, against VOC in SLE patients to levels comparable to healthy controls.

**Figure 2.**
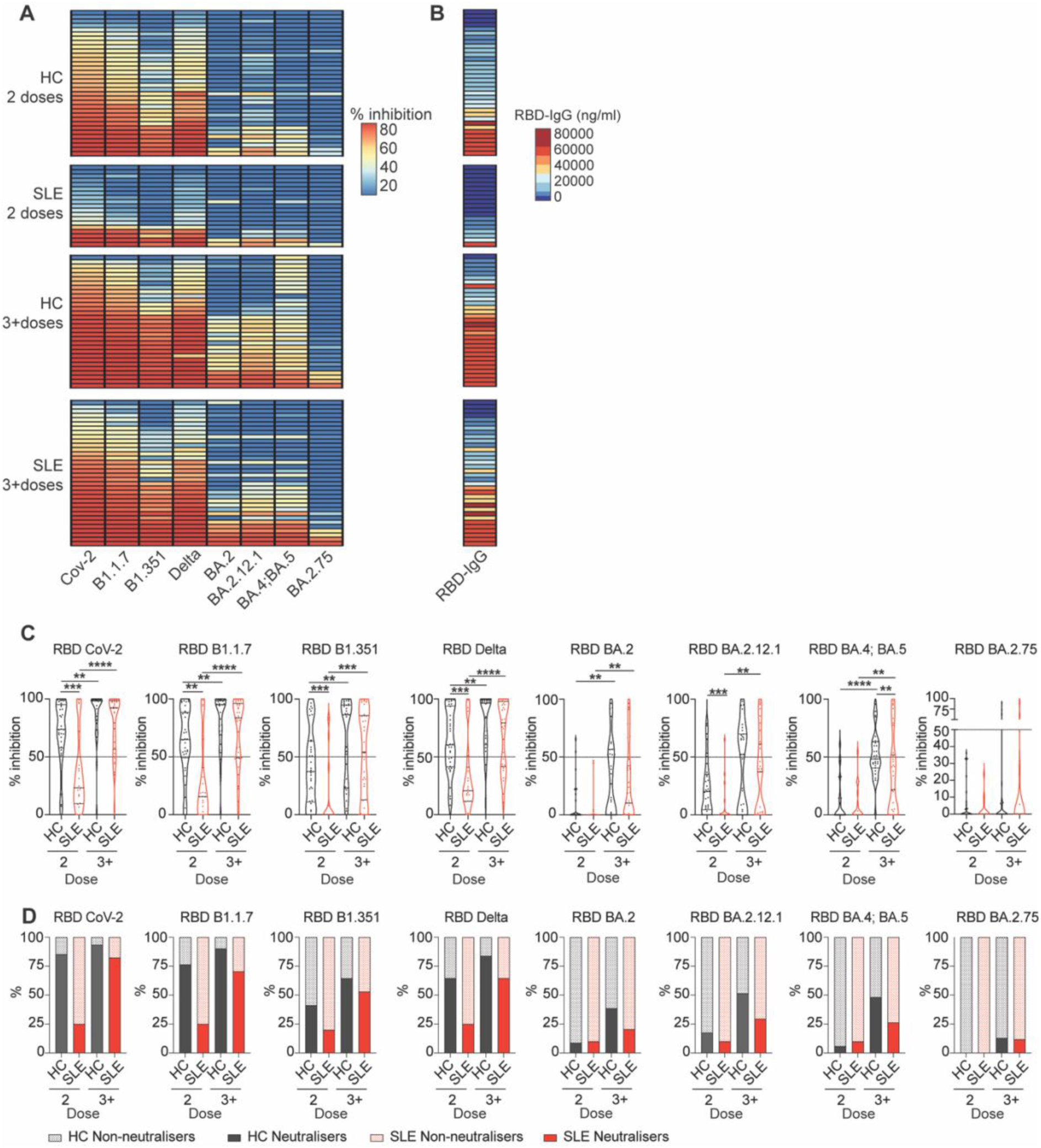
SLE patients show reduced neutralising capacity against SARS-CoV-2 variants of concern after initial vaccination. **(A)** Heatmap showing percentage inhibition values against the RBD of different SARS-CoV-2 strains (CoV-2, B1.1.7, B1.351, Delta, BA.2, BA.2.12.1, BA.4;BA.5, and BA.2.75) of serum isolated from a cross-sectional cohort of SLE patients and healthy controls (HC), after 2 or 3^+^ doses of vaccine. **(B)** Heatmaps showing the titres of RBD-specific IgG for the corresponding individuals in (A). **(C)** Cumulative data violin plots showing percentage inhibition values against all tested different SARS-CoV-2 strains (CoV-2, B1.1.7, B1.351, Delta, BA.2, BA.2.12.1, BA.4;BA.5, and BA.2.75) of serum isolated from a cross-sectional cohort of SLE patients and healthy controls (HC), after 2 or 3^+^ doses of vaccine. Dotted line on graph indicates the neutralisation threshold (50%). **(D)** Stacked bar plots showing the percentage of neutralisers for all tested different SARS-CoV-2 strains (CoV-2, B1.1.7, B1.351, Delta, BA.2, BA.2.12.1, BA.4;BA.5, and BA.2.75) within each group. \*\**P*<0.01, \*\*\**P*<0.001, \*\*\*\**P*<0.0001, by (C) by Mann-Whitney of selected group pairs or by Wilcoxon matched-pairs signed rank test. Violin plots display the median, along with the 25th and 75th percentiles.

### SARS-CoV-2-specific B cell isotype switching is impaired in SLE patients

SLE patients present with several inherent B cell abnormalities including increases in transitional-2, DN B cells, and plasma cells (17–19). We found increased levels of B cells after 2 and 3^+^doses in SLE patients compared to healthy individuals (Figure 3A). We report decreased levels of total memory B cells (MBC; CD27^+^IgD^-^CD24^+^CD38^lo^) and double positive B cells (DP; CD27^+^IgD^+^), mirrored by an increased in double negative B cells (DN; CD27^-^IgD^-^) and plasmablasts (CD27^+^IgD^-^CD24^-^CD38^+^) in SLE patients compared to healthy controls at baseline, differences that were maintained after vaccination (Figure 3A; gating strategy in supplemental Figure 4A,B).

**Figure 3.**
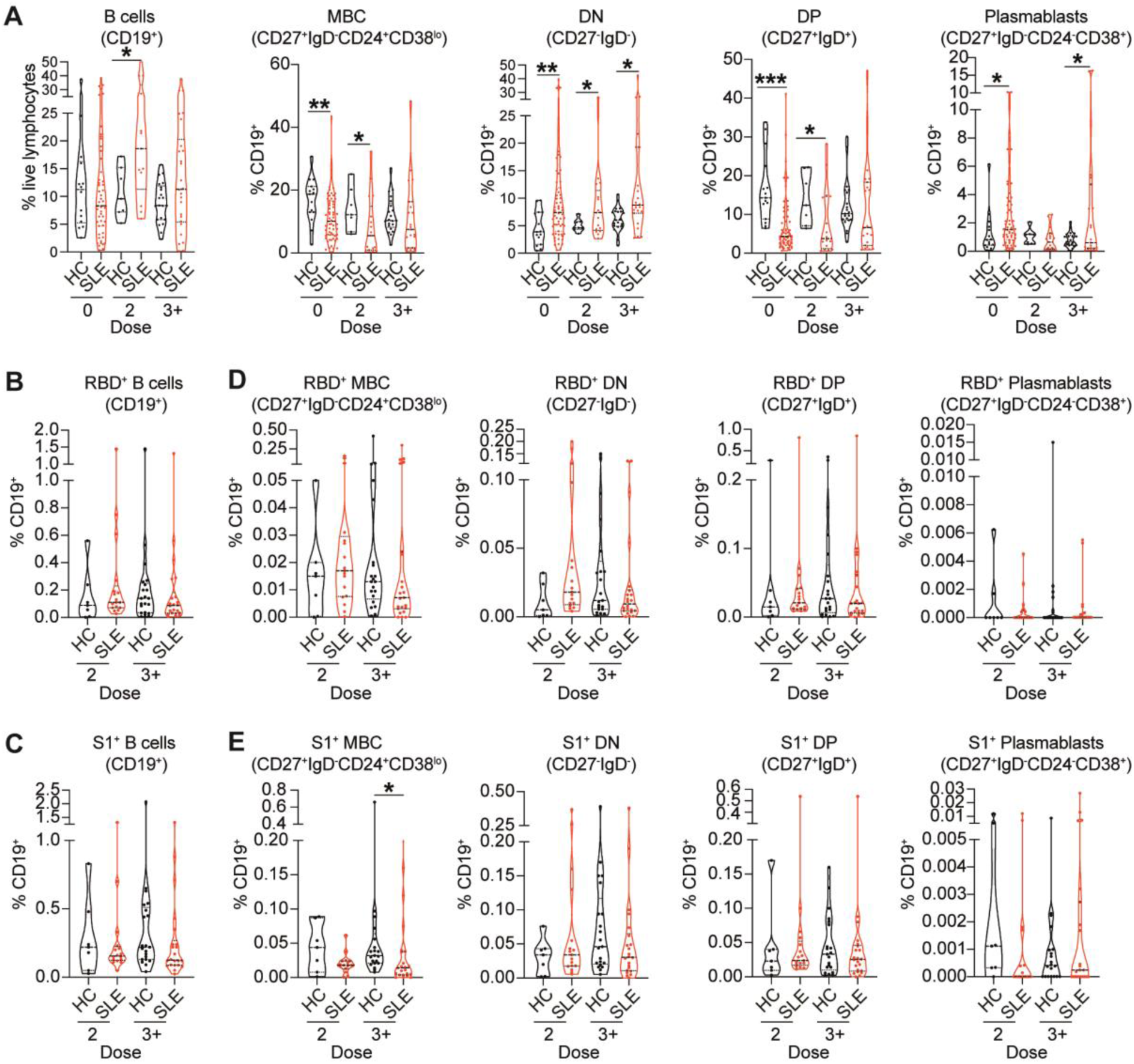
SLE patients show no differences in the frequencies of SARS-CoV-2-specific B cell subsets compared to healthy individuals. **(A)** Cumulative data violin plots showing frequencies of total B cells (CD19^+^), memory B cells (MBC, CD27^+^IgD^-^CD24^+^CD38^lo^), double-negative B cells (DN, CD27^-^IgD^-^), double-positive B cells (DP, CD27^+^IgD^+^) and plasmablasts (CD27^+^IgD^-^CD24^-^CD38^+^) within the total CD19^+^B cell population in *ex vivo* peripheral blood of healthy controls (HC) and SLE patients, pre-vaccination (0 dose) or after 2 or 3^+^ doses of vaccine. **(B,C)** Cumulative data violin plots showing frequencies of total RBD^+^B cells (B) and S1^+^B cells (C) in *ex vivo* peripheral blood of healthy controls (HC) and SLE patients, pre-vaccination (0 dose) or after 2 or 3^+^ doses of vaccine. **(D,E)** Cumulative data violin plots showing frequencies of RBD^+^ (D) and S1^+^ (E) MBC, DN B cells, DP B cells and plasmablasts, within the total CD19^+^B cell population in *ex vivo* peripheral blood of HC and SLE patients, pre-vaccination (0 dose), or after 2 or 3^+^ doses of vaccine. (\**P*<0.05, \*\**P*<0.01 \*\*\**P*<0.001, \*\*\*\**P*<0.0001, by unpaired t-test, Mann-Whitney test or data transformation according to data distribution and features (see methods). Violin plots display the median, along with the 25th and 75th percentiles.

To understand whether changes in B cell subsets frequency observed in SLE patients, impacted the response to SARS-CoV-2 vaccination, we measured RBD and S1-specific memory B cells, DN, DP, and plasmablasts frequencies in SLE patients and healthy controls (demographics in supplemental table 3; gating strategy in supplemental Figure 4A,B). There was no difference in the frequencies of total RBD^+^ and S1^+^ B cells between SLE patients and healthy controls (Figure 3B,C). Analysis of antigen-specific B cell subsets, amongst total B cells, show that the frequencies of RBD^+^ and S1^+^MBCs in SLE patients were decreased (a trend for RBD and significant for S1) compared to healthy controls even after 3^+^ doses of vaccine (Figure 3D,E). We did not observe any differences between healthy controls and SLE patients in the frequencies of RBD^+^ or S1^+^DN B cells, DP B cells or plasmablasts (Figure 3D,E).

Next, we assessed how repeated vaccinations affects the frequencies of memory unswitched (IgM^+^) or memory switched (IgG^+^/IgA^+^) RBD^+^ and S1^+^ B cells (Figure 4A). There was an increased frequency of RBD^+^/S1^+^ IgM^+^unswitched MBCs following 2 and 3^+^ doses in SLE patients (Figure 4B,C), and an increase in and RBD^+^/S1^+^ IgM^+^ DN B cells after 2 doses (supplemental Figure 4C,D). This was accompanied by a decrease in RBD^+^/S1^+^ IgG^+^ MBCs (Figure 4D,E), and RBD^+^/S1^+^ IgG^+^ DN B cells (supplemental Figure 4C,D) after 2 or 3^+^ doses in SLE patients compared to healthy controls. We also reported increased frequencies of RBD^+^/S1^+^ IgM^+^ early plasmablasts (early-PB) (CD27^+^IgD^-^CD24^mid^CD38^+^) in SLE patients compared to healthy controls (supplemental Figure 4E,F), previously shown to arise from extrafollicular responses (17–19).

**Figure 4.**
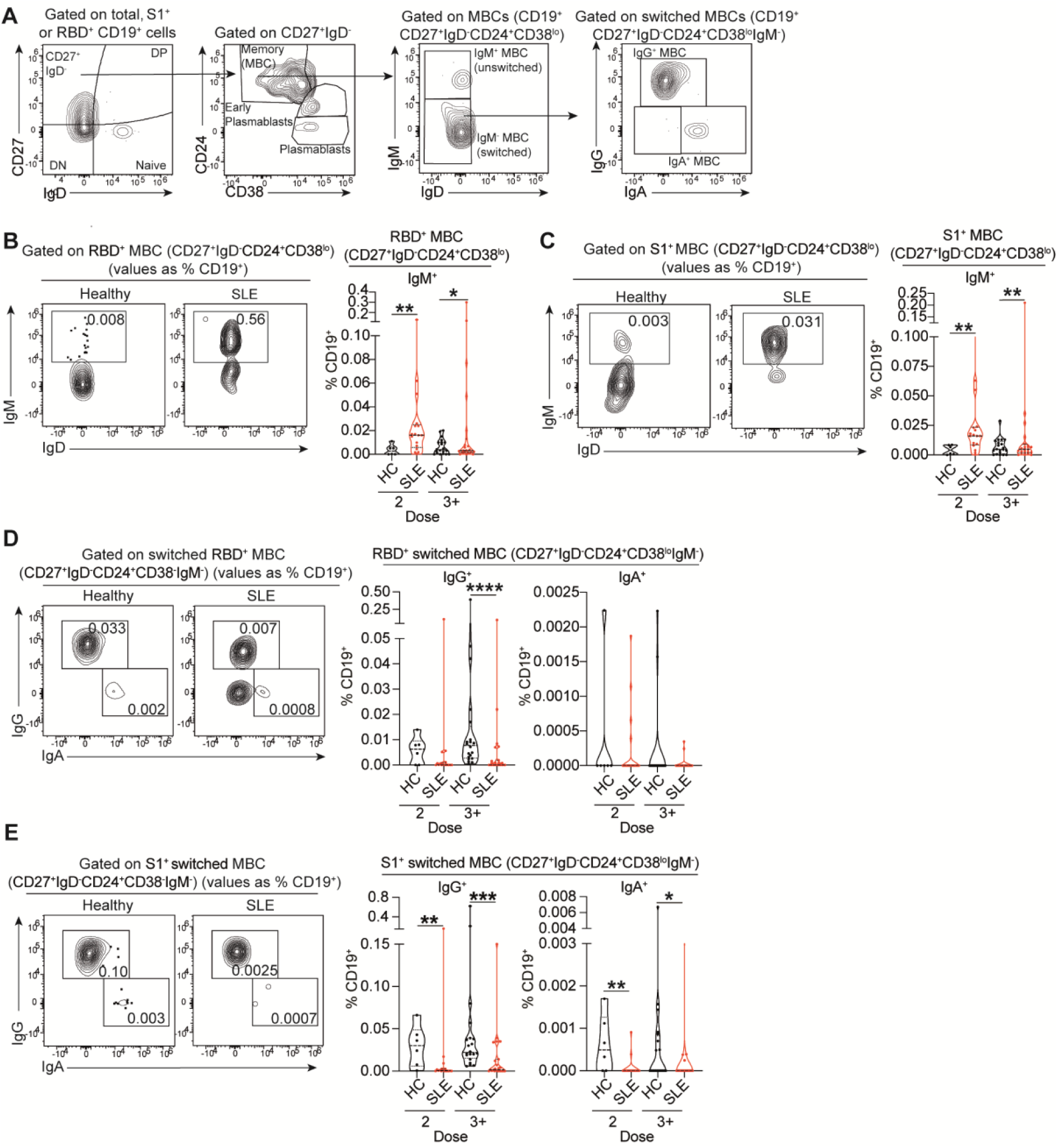
SARS-CoV-2-specific B cells show lower IgG and IgA class-switching in SLE patients. **(A)** Representative contour plots showing the gating strategy for identification of memory B cells (MBCs; CD27^+^IgD^-^CD24^+^CD38^lo^), early plasmablasts (CD27^+^IgD^-^ CD24^mid^CD38^+^), and plasmablasts (CD27^+^IgD^-^CD24^-^CD38^hi^), within RBD^+^ and S1^+^ B cell subsets. IgM^+^ unswitched (CD27^+^IgD^-^CD24^+^CD38^lo^IgM^+^) and IgG^+^ switched (CD27^+^IgD^-^CD24^+^CD38^lo^IgM^-^IgG^+^) or IgA^+^ switched (CD27^+^IgD^-^CD24^+^CD38^lo^IgM^-^ IgA^+^) MBCs were identified within RBD^+^ and S1^+^ B cell subsets. **(B,C)** Representative contour plots and cumulative data violin plots showing the frequencies of (B) RBD-specific and (C) S1-specific IgM^+^ unswitched MBCs (CD27^+^IgD^-^CD24^+^CD38^lo^IgM^+^) in *ex vivo* peripheral blood of healthy controls (HC) and SLE patients, after 2 or 3^+^ doses of vaccine. Values in gates indicate the percentage within CD19^+^ cells. **(D,E)** Representative contour plots and cumulative data violin plots showing the frequencies of (D) RBD-specific and (E) S1-specific IgG^+^ and IgA^+^ switched MBCs (CD27^+^IgD^-^ CD24^+^CD38^lo^IgM^-^ IgG^+^/IgA^+^), in *ex vivo* peripheral blood of healthy controls (HC) and SLE patients after 2 or 3^+^ doses of vaccine. Values in gates indicate the percentage within CD19^+^ cells. \**P*<0.05, \*\**P*<0.01 \*\*\**P*<0.001, \*\*\*\**P*<0.0001, by Mann-Whitney test or data transformation according to data distribution and features (see methods). Violin plots display the median, along with the 25th and 75th percentiles.

The results for RBD^+^IgA^+^ MBCs were difficult to interpret due to very low frequencies of RBD^+^IgA^+^ B cells (Figure 4D). Nevertheless, S1^+^IgA^+^MBCs were significantly reduced in SLE patients after 2 and 3^+^ doses, compared to healthy controls (Figure 4E). No differences were observed between SLE patients and healthy controls in the frequencies of IgA^+^DN B cells for both RBD^+^ /S1^+^ and populations (supplemental Figure 4C,D).

The accumulation of antigen-specific IgM^+^ early-PB in SLE patients suggest a defect in CSR supported by an extrafollicular differentiation pathway. We calculated the ratio between RBD^+^/S1^+^ switched (IgA or IgG) MBCs and RBD^+^/S1^+^ IgM^+^ early-PB as an index (CSR ratio) of efficient CSR compared to extrafollicular responses. SLE patients showed a significantly lower CSR ratio compared to healthy controls (supplemental Figure 5A,B). A similar CSR impairment was also observed in non-antigen-specific (S1^-^) B cells (supplemental Figure 5C-F), suggesting that this defect may not be limited to vaccine-derived antigen-specific B cells.

### IFNα impairs class-switch recombination *in vitro* and promotes IgM^+^ early plasmablasts

Our findings and those from others (20) suggest that SLE patients exhibit impaired CSR and increased frequencies of vaccine-specific IgM^+^ MBCs and IgM^+^ early-PBs. Given the pivotal role of IFNα in SLE pathogenesis and in driving plasma cell differentiation (21,22), we hypothesised that exposure to elevated IFNα levels may contribute to the observed CSR imbalance in SLE patients.

We took advantage of a previously established culture system whereby healthy naive B cells were stimulated *in vitro* for 6 days with a combination of anti-IgM, CD40L, and IFNγ to induce class-switching to IgG isotypes (23). Increasing concentrations of IFNα were added to the culture system to model the potential effect of IFNα on B cell differentiation and CSR of SLE B cells (Figure 5A). As expected, IFNα reduced memory B cells whilst promoting an increase of plasmablasts *in vitro* (Figure 5B,C). In addition, high IFNα levels reduced switching of healthy naive B cells to IgG1, IgG2 and IgG3, under CSR-IgG-polarising conditions (Figure 5D,E). Supporting our *ex vivo* findings in SLE patient B cells, we observed a significant increase of IgM^+^ early-PB frequencies and a decreased ratio of IgG^+^ B cells to IgM^+^ early-PBs (CSR ratio) under these polarizing *in vitro* conditions (Figure 5F-H). Together this *in vitro* modelling of SLE B cell CSR indicates that high concentrations of circulating IFNα in SLE patients may contribute to the impaired CSR of B cells and subsequently result in weaker antibody responses following SARS-CoV-2 vaccination.

**Figure 5.**
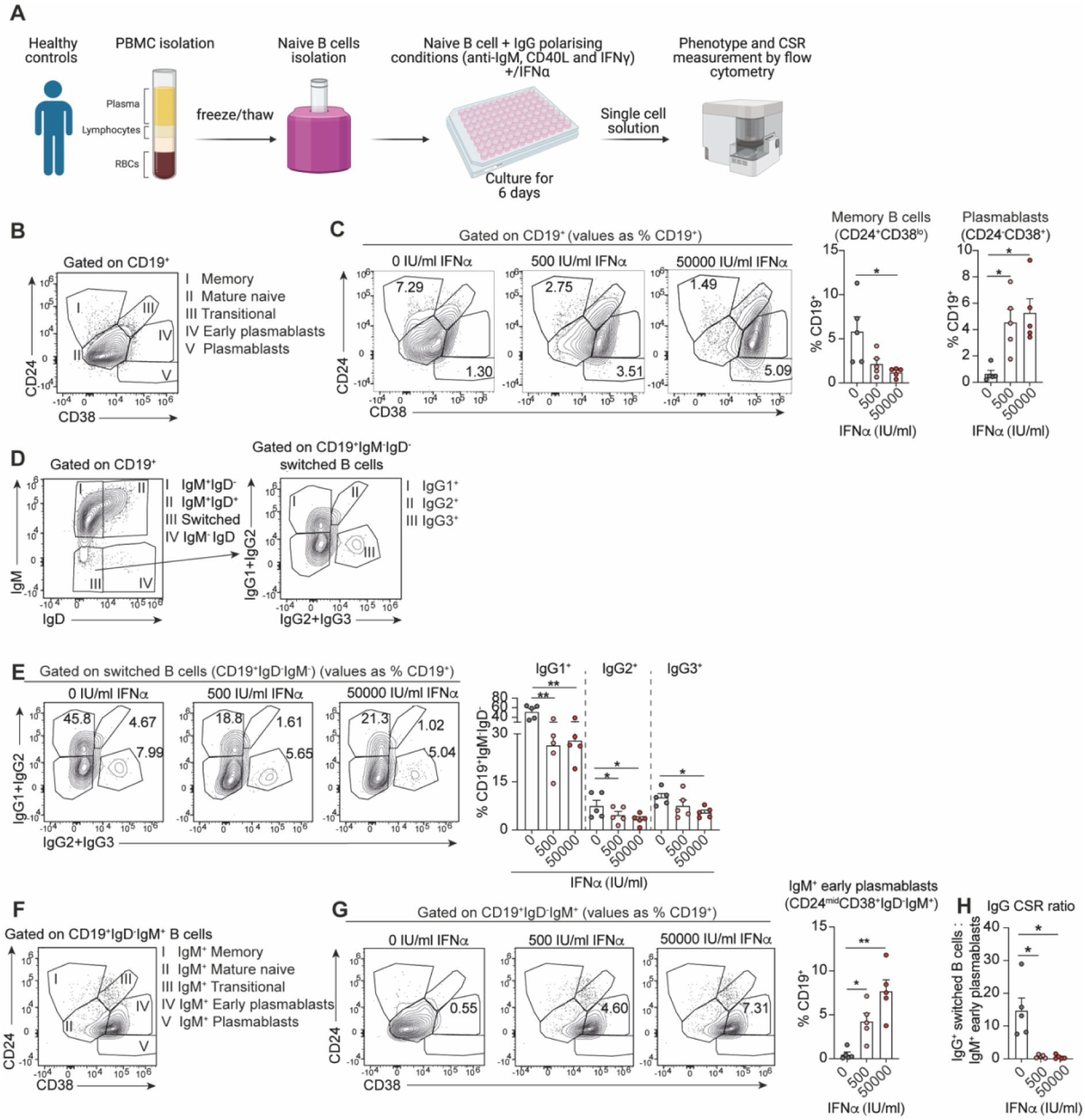
Interferon alpha suppresses healthy naive B cell class switching to IgG. **(A)** Schematic of class-switching *in vitro* experiment. Naive B cells (CD27^-^IgD^+^) were isolated from previously frozen PBMCs of healthy controls. Isolated naive B cells were cultured under IgG class switching polarising conditions (anti-IgM, CD40L and IFNγ) together with 0, 500, or 50000 IU/ml of IFNα for 6 days. After 6 days of culture, cells were harvested and stained for flow cytometry to assess their phenotype and CSR status. **(B)** Contour plots showing the gating strategy for the identification of memory B cells (CD19^+^CD24^+^CD38^lo^), mature naive B cells (CD19^+^CD24^mid^CD38^mid^), transitional B cells (CD19^+^CD24^+^CD38^+^), early plasmablasts (CD19^+^CD24^mid^CD38^+^) and plasmablasts (CD19^+^CD24^-^CD38^+^) after 6 days in IgG-polarising class-switch stimulation of isolated healthy naive B cells (CD27^-^IgD^+^). **(C)** Representative contour plots and cumulative data bar graphs showing the frequencies of memory B cells (CD19^+^CD24^+^CD38^lo^) and plasmablasts (CD19^+^CD24^-^CD38^+^) after 6 days stimulation of isolated healthy naive B cells under IgG-polarising conditions and increasing concentrations of recombinant IFNα. Values in gates indicate the percentage within CD19^+^ cells. **(D)** Contour plots showing the gating strategy for the identification of IgG1^+^, IgG2^+^ and IgG3^+^ switched B cells after 6 days stimulation of isolated healthy naive B cells under IgG-polarising conditions. **(E)** Representative contour plots and cumulative bar graphs showing the frequencies of IgG1^+^, IgG2^+^ and IgG3^+^ switched B cells, after 6 days stimulation of isolated healthy naive B cells under IgG-polarising conditions and increasing concentrations of recombinant IFNα. Values in gates indicate the percentage within CD19^+^IgD^-^IgM^-^ cells. **(F)** Contour plots showing the gating strategy for the identification of IgM^+^ memory, IgM^+^ mature naive, IgM^+^ transitional B cells, IgM^+^ early plasmablasts and IgM^+^ plasmablasts after 6 days in IgG-polarising class-switch stimulation of isolated healthy naive B cells. **(G)** Representative contour plots and cumulative bar graphs showing the frequencies of IgM^+^ early plasmablasts (CD19^+^CD24^mid^CD38^+^ IgD^-^IgM^+^) after 6 days stimulation of isolated healthy naive B cells under IgG-polarising conditions and increasing concentrations of recombinant IFNα. Values in gates indicate the percentage within CD19^+^ cells. **(H)** Cumulative data bar graph showing the ratio between IgG^+^ switched memory B cells (sum of IgG1^+^, IgG2^+^ and IgG3^+^ frequencies within CD19^+^IgD^-^IgM^-^ cells) and IgM^+^ early plasmablasts, after 6 days stimulation of isolated healthy naive B cells under IgG-polarising conditions and increasing concentrations recombinant IFNα. \**P*<0.05, \*\**P*<0.01, by Kruskal-Wallis test with Dunn’s multiple comparison. Error bars in bar plots show mean+/-SEM.

## DISCUSSION

The efficacy of the SARS-CoV-2 vaccination programme has been well-documented in non-immunocompromised cohorts, however our understanding of how SLE patients respond to new vaccines remains unknown. Here we show that SLE patients failed to generate robust SARS-CoV-2-specific protective antibody responses after 2 doses, including vaccine-specific IgG and IgA, and neutralising capacity against the original strain of the virus and VoC. We demonstrate that booster vaccination provides sufficient protection in around three quarters of SLE patients compared to the levels of protection reached in healthy individuals. Others have reported similar findings, as two-thirds of SLE patients treated with belimumab seroconverted only after booster vaccination (24). Booster vaccination also improved RBD-IgG responses in an adolescent SLE cohort, and enabled seroconversion against Omicron variants (25). It should be noted that even with booster vaccination many SLE patients did not acquire sufficient humoral protection, particularly against several VoC. These observations highlight the need for tailored vaccination strategies for SLE patients and offer important lessons for future pandemics.

The reduced humoral protection observed in SLE patients after initial SARS-CoV-2 vaccination has been previously linked to immunosuppressant drugs (7,26). However, there is no clear consensus on whether specific SLE treatments impair vaccine efficacy, and current research (27), including our study, shows variable findings. The impaired efficacy of SARS-CoV-2 vaccination may also be the consequence of SLE disease activity, as we observed higher RBD-IgM responses in patients with active disease. Nevertheless, the variability observed between patients suggests that multiple factors may contribute to the reduced vaccine responses in SLE patients.

We provide novel insight by reporting that vaccine-specific memory and DN B cells in SLE patients express IgM without class-switching to IgG or IgA. Additionally, SLE patients present with increased frequencies of vaccine-specific IgM^+^ early plasmablasts. It is tempting to speculate that SARS-CoV-2 vaccine responses in SLE patients occur extrafollicularly. As extrafollicular reactions typically have lower CSR levels and rapid IgM^+^ plasmablast generation compared to follicular and GC-dependent B cell responses (28). It has been suggested that extrafollicular reactions provide reduced vaccine efficacy. Indeed, expansions of DN B cells have been linked to reduced SARS-CoV-2 neutralising responses and lower frequencies of RBD-specific B cells (29). Notably, extrafollicular DN B cells highly express autoimmune regulator (AIRE), and a recent preprint demonstrated that AIRE in GC B cells suppresses AID-mediated B cell affinity maturation and CSR (30). Whether AIRE expression in extrafollicular B cell responses contributes to the impaired CSR observed in SLE patients will be subject of our future investigation.

Finally, we demonstrated that *in vitro* stimulation of healthy B cells with IFNα recapitulated the reduced IgG CSR and expanded IgM^+^ early plasmablast population seen in SLE patients *ex vivo*. IFNα may contribute to reduced SARS-CoV-2 vaccine responses in SLE patients. High levels of serum IFNα and upregulation of interferon-inducible genes are associated with active SLE (21,22), and we identified that clinically active patients present with increased RBD-IgM responses compared to inactive patients. IFNα signalling is also shown to promote extrafollicular B cell reactions (28,31,32). The exact impact of IFNα on SARS-CoV-2 vaccine responses in SLE patients remains unclear and warrants further study.

There are limitations to this study. Our cohort of longitudinal SLE patients was small, as the majority of SLE patients were shielding during the initial vaccination programme. We were also only able to analyse a small number of patients treated with DMARDs, and other drugs included in the study, which prevent us from drawing conclusions about the potential effect of individual medications or dosage on vaccine efficacy.

In summary we report that although repeated doses of vaccine improve protective responses in SLE patients, they remain significantly lower than those achieved in healthy individuals. We also report a skewed memory B cells formation towards an accumulation of unswitched IgM^+^ memory B cells. As such, continued booster and optimisation of vaccination strategy should be considered for patients with SLE.

## Data Availability

All data produced in the present study are available upon reasonable request to the authors

## DATA AVAILABILITY STATEMENT

Data are available upon request.

## ETHICS STATEMENTS

Consent obtained directly from patients. This study involves human participants and was obtained from the UCLH Health Service Trust ethics committee, under REC reference no. 14/SC/1200. Patients gave informed consent to participate in the study before taking part.

## ACKNOWLEDGEMENTS

We thank Dr Thomas McDonnell for guidance with experimental approaches and feedback. This work was presented at: British Society of Immunology (BSI) Focus B cell meeting in April 2022 in London (United Kingdom), and at the BSI Annual Congress in December 2022 in Liverpool (United Kingdom).

## AUTHOR CONTRIBUTIONS

All authors were involved in drafting the article or revising it critically for important intellectual content, and all authors approved the final version to be published. Professor Mauri had full access to all data in the study and takes responsibility for the integrity of the data and the accuracy of the data analysis.

**Study conception and design**

Mauri, Montamat, Bradford.

**Acquisition of data**

Montamat, Meehan, Yıldırım, Guimarães, Goldblatt, Johnson, Isenberg.

**Analysis and interpretation of data**

Mauri, Montamat, Meehan, Bradford.

## Funding

This study was funded by a LUPUS UK grant (185431), (186075) (186987) and Versus Arthritis (17603) awarded to C.Mauri.

## Competing interests

None declared.

## Patient and public involvement

Patients and/or the public were not involved in the design or conduct, reporting, or dissemination plans of this research.

## Notes

### Competing Interest Statement

The authors have declared no competing interest.

### Author Declarations

This study involves human participants and was obtained from the UCLH Health Service Trust ethics committee, under REC reference no. 14/SC/1200. Patients gave informed consent to participate in the study before taking part.

## REFERENCES

1. Hurtado C, Rojas-Gualdrón DF, Urrego R, Cashman K, Vásquez-Trespalacios EM, Díaz-Coronado JC, et al. Altered B cell phenotype and CD27+ memory B cells are associated with clinical features and environmental exposure in Colombian systemic lupus erythematosus patients. Front Med (Lausanne) 2022;9.

2. Mauri C, Menon M. Human regulatory B cells in health and disease: Therapeutic potential. Journal of Clinical Investigation 2017;127.

3. Jones DD, Wilmore JR, Allman D. Cellular Dynamics of Memory B Cell Populations: IgM+ and IgG+ Memory B Cells Persist Indefinitely as Quiescent Cells. The Journal of Immunology 2015;195.

4. Mesin L, Ersching J, Victora GD. Germinal Center B Cell Dynamics. Immunity 2016;45.

5. Roco JA, Mesin L, Binder SC, Nefzger C, Gonzalez-Figueroa P, Canete PF, et al. Class-Switch Recombination Occurs Infrequently in Germinal Centers. Immunity 2019;51.

6. Laidlaw BJ, Ellebedy AH. The germinal centre B cell response to SARS-CoV-2. Nat Rev Immunol 2022;22.

7. Petri M, Joyce D, Haag K, Fava A, Goldman DW, Zhong D, et al. Effect of Systemic Lupus Erythematosus and Immunosuppressive Agents on COVID-19 Vaccination Antibody Response. Arthritis Care Res (Hoboken) 2023;75.

8. Yuki EFN, Borba EF, Pasoto SG, Seguro LP, Lopes M, Saad CGS, et al. Impact of Distinct Therapies on Antibody Response to SARS-CoV-2 Vaccine in Systemic Lupus Erythematosus. Arthritis Care Res (Hoboken) 2022;74.

9. Izmirly PM, Kim MY, Samanovic M, Fernandez-Ruiz R, Ohana S, Deonaraine KK, et al. Evaluation of Immune Response and Disease Status in Systemic Lupus Erythematosus Patients Following SARS–CoV-2 Vaccination. Arthritis and Rheumatology 2022;74.

10. Sjöwall J, Azharuddin M, Frodlund M, Zhang Y, Sandner L, Dahle C, et al. SARS-CoV-2 Antibody Isotypes in Systemic Lupus Erythematosus Patients Prior to Vaccination: Associations With Disease Activity, Antinuclear Antibodies, and Immunomodulatory Drugs During the First Year of the Pandemic. Front Immunol 2021;12.

11. Moyon Q, Sterlin D, Miyara M, Anna F, Mathian A, Lhote R, et al. BNT162b2 vaccine-induced humoral and cellular responses against SARS-CoV-2 variants in systemic lupus erythematosus. Ann Rheum Dis 2022;81.

12. Ammitzbøll C, Bartels LE, Bøgh Andersen J, Risbøl Vils S, Elbæk Mistegård C, Dahl Johannsen A, et al. Impaired Antibody Response to the BNT162b2 Messenger RNA Coronavirus Disease 2019 Vaccine in Patients With Systemic Lupus Erythematosus and Rheumatoid Arthritis. ACR Open Rheumatol 2021;3.

13. Amanat F, Stadlbauer D, Strohmeier S, Nguyen THO, Chromikova V, McMahon M, et al. A serological assay to detect SARS-CoV-2 seroconversion in humans. Nat Med 2020;26.

14. Cuadrado MJ, Calatayud I, Urquizu-Padilla M, Wijetilleka S, Kiani-Alikhan S, Karim MY. Immunoglobulin abnormalities are frequent in patients with lupus nephritis. BMC Rheumatol 2019;3.

15. Lim E, Tao Y, White AJ, French AR, Cooper MA. Hypogammaglobulinemia in pediatric systemic lupus erythematosus. Lupus 2013;22.

16. Tan CY, Chiew CJ, Pang D, Lee VJ, Ong B, Lye DC, et al. Protective immunity of SARS-CoV-2 infection and vaccines against medically attended symptomatic omicron BA.4, BA.5, and XBB reinfections in Singapore: a national cohort study. Lancet Infect Dis 2023;23.

17. Karrar S, Cunninghame Graham DS. Abnormal B Cell Development in Systemic Lupus Erythematosus: What the Genetics Tell Us. Arthritis and Rheumatology 2018;70.

18. Iwata S, Tanaka Y. B-cell subsets, signaling and their roles in secretion of autoantibodies. Lupus 2016;25.

19. Jenks SA, Wei C, Bugrovsky R, Hill A, Wang X, Rossi FM, et al. B cell subset composition segments clinically and serologically distinct groups in chronic cutaneous lupus erythematosus. Ann Rheum Dis 2021;80.

20. Faliti CE, Anam FA, Cheedarla N, Woodruff MC, Usman SY, Runnstrom MC, et al. Poor immunogenicity upon SARS-CoV-2 mRNA vaccinations in autoimmune SLE patients is associated with pronounced EF-mediated responses and anti-BAFF/Belimumab treatment. medRxiv 2023.

21. Bradford HF, Haljasmägi L, Menon M, McDonnell TCR, Särekannu K, Vanker M, et al. Inactive disease in patients with lupus is linked to autoantibodies to type I interferons that normalize blood IFNα and B cell subsets. Cell Rep Med 2023;4.

22. Menon M, Blair PA, Isenberg DA, Mauri C. A Regulatory Feedback between Plasmacytoid Dendritic Cells and Regulatory B Cells Is Aberrant in Systemic Lupus Erythematosus. Immunity 2016;44.

23. Ng JCF, Montamat Garcia G, Stewart AT, Blair P, Mauri C, Dunn-Walters DK, et al. sciCSR infers B cell state transition and predicts class-switch recombination dynamics using single-cell transcriptomic data. Nat Methods 2024;21.

24. Tunitsky-Lifshitz Y, Maoz-Segal R, Niznik S, Shavit R, Haj Yahia S, Langevitz P, et al. The third dose of BNT162b2 COVID-19 vaccine is efficacious and safe for systemic lupus erythematosus patients receiving belimumab. Lupus 2023;32.

25. Piyaphanee N, Charuvanij S, Thepveera S, Toh ZQ, Licciardi P V., Pattaragarn A, et al. Immunogenicity and safety of BNT162b2 vaccination in adolescents with systemic lupus erythematosus. Lupus 2024;33.

26. Garcia-Cirera S, Calvet J, Berenguer-Llergo A, Pradenas E, Marfil S, Massanella M, et al. Glucocorticoids’ treatment impairs the medium-term immunogenic response to SARS-CoV-2 mRNA vaccines in Systemic Lupus Erythematosus patients. Sci Rep 2022;12.

27. Saxena A, Guttmann A, Masson M, Kim MY, Haberman RH, Castillo R, et al. Evaluation of SARS-CoV-2 IgG antibody reactivity in patients with systemic lupus erythematosus: analysis of a multi-racial and multi-ethnic cohort. Lancet Rheumatol 2021;3.

28. Al-Aubodah TA, Aoudjit L, Pascale G, Perinpanayagam MA, Langlais D, Bitzan M, et al. The extrafollicular B cell response is a hallmark of childhood idiopathic nephrotic syndrome. Nat Commun 2023;14.

29. Yam-Puc JC, Hosseini Z, Horner EC, Gerber PP, Beristain-Covarrubias N, Hughes R, et al. Age-associated B cells predict impaired humoral immunity after COVID-19 vaccination in patients receiving immune checkpoint blockade. Nat Commun 2023;14.

30. Zhou JZ, Huang B, Pei B, Sun GW, Pawlitz MD, Zhang W, et al. A Germinal Center Checkpoint of AIRE in B Cells Limits Antibody Diversification. bioRxiv 2024.

31. Peng SL, Szabo SJ, Glimcher LH. T-bet regulates IgG class switching and pathogenic autoantibody production. Proc Natl Acad Sci U S A 2002;99.

32. Swanson CL, Wilson TJ, Strauch P, Colonna M, Pelanda R, Torres RM. Type I IFN enhances follicular B cell contribution to the T cell-independent antibody response. Journal of Experimental Medicine 2010;207.

