## Supplemental material for "Reduced response to SARS-CoV-2 vaccination is associated with impaired immunoglobulin class switch recombination in SLE patients"

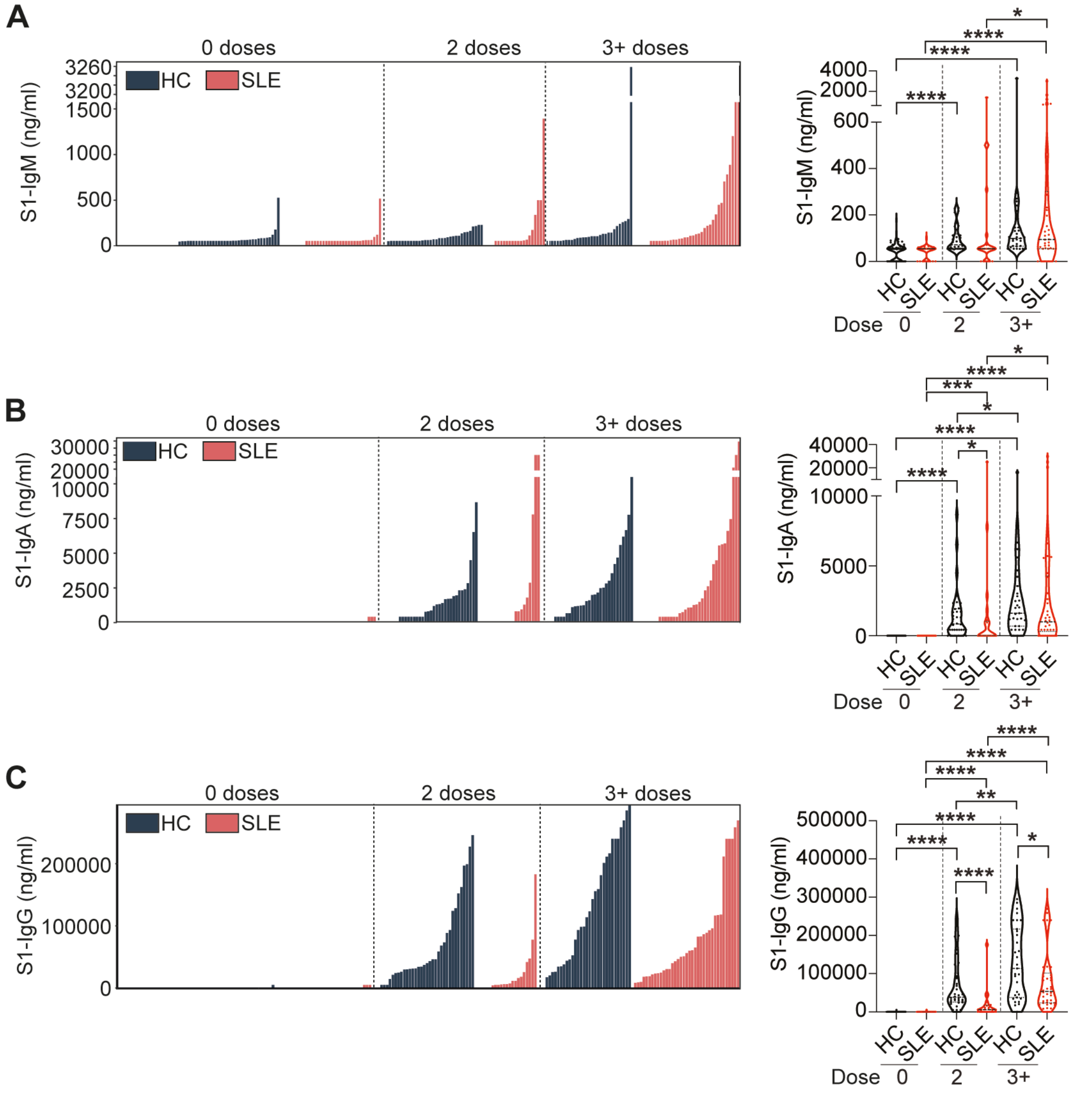


**Supplementary Figure 1.**

**(A-C)** Individual data bar plots and cumulative data violin plots showing the titres of S1-specific (A) IgM, (B) IgA, and (C) IgG antibodies in a cross-sectional cohort of SLE patients and healthy controls (HC), pre-vaccination (0 doses) and following 2 or $3^+$ doses of vaccine. *$P$<0.05, **$P$<0.01, ***$P$<0.001, ****$P$<0.0001, by Mann-Whitney test of selected group pairs. Violin plots display the median, along with the 25th and 75th percentiles.

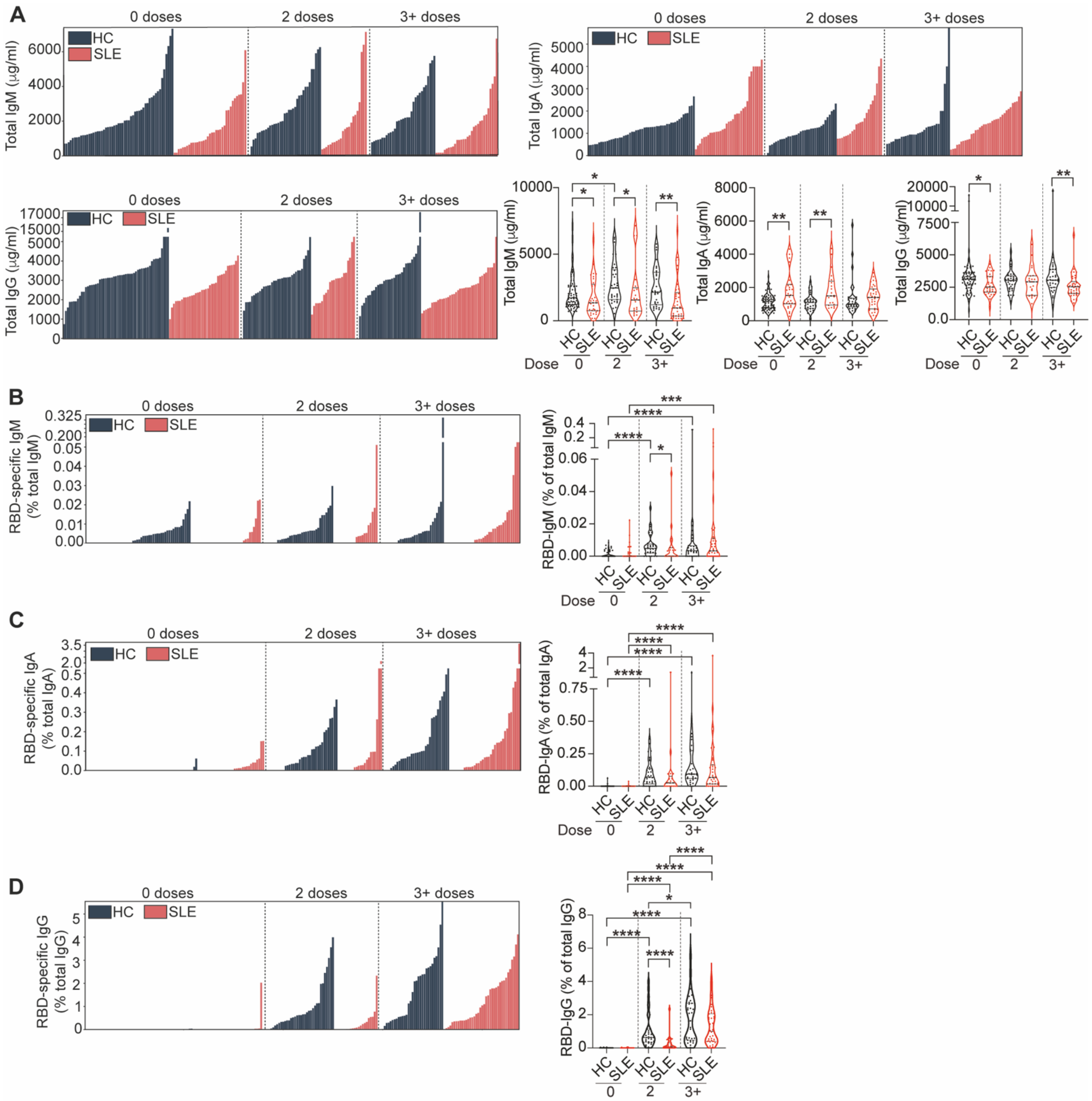

**Supplementary Figure 2.**

**(A)** Individual data bar plots and cumulative data violin plots showing the titres of total IgM, IgA and IgG antibodies in a cross-sectional cohort of SLE patients and healthy controls (HC), pre-vaccination (0 doses) and following 2 or 3$^{+}$ doses of vaccine. **(B-D)** Individual bar plots and cumulative data violin plots showing the titres of (B) RBD-specific IgM antibodies as a percentage of total IgM, (C) RBD-specific IgA antibodies as a percentage of total IgA, and (D) RBD-specific IgG antibodies as a percentage of total IgG in a cross-sectional cohort of SLE patients and HC, pre-vaccination and following 2 or 3$^{+}$ doses of vaccine. *$P$<0.05, **$P$<0.01, ***$P$<0.001, ****$P$<0.0001, by Mann-Whitney test of selected group pairs. Violin plots display the median, along with the 25th and 75th percentiles.

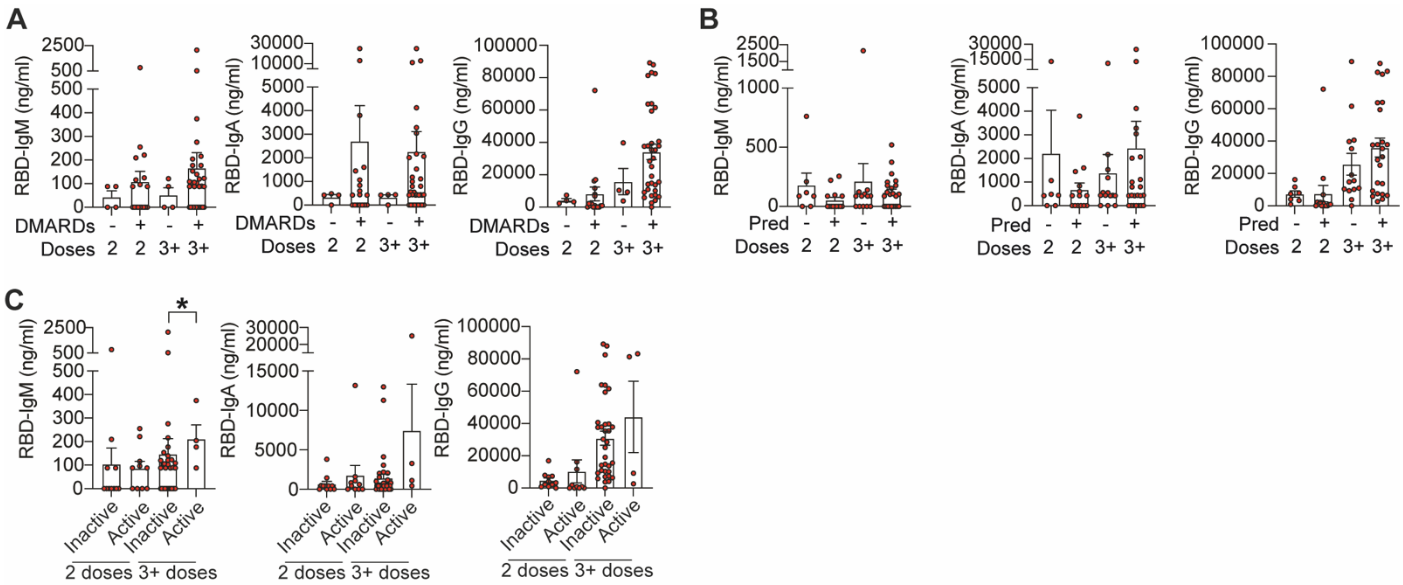


**Supplementary Figure 3.**

**(A)** Cumulative data bar plots showing the titres of RBD-specific IgM, IgA and IgG antibodies in SLE patients treated with or without disease-modifying anti-rheumatic drugs (DMARDs, including hydroxychloroquine, mycophenolate mofetil, methotrexate and azathioprine), after 2 or $3^{+}$ doses of vaccine. **(B)** Cumulative data bar plots showing the titres of RBD-specific IgM, IgA and IgG antibodies in SLE patients treated with or without prednisolone, after 2 or $3^{+}$ doses of vaccine. **(C)** Cumulative data bar plots showing the titres of RBD-specific IgM, IgA and IgG antibodies in SLE patients with active (BILAG score ≥ 5) or inactive disease (BILAG score < 5), after 2 or $3^{+}$ doses of vaccine. **P*<0.05, ***P*<0.01, ****P*<0.001, by (A to C) by Mann-Whitney test of selected group pairs. Error bars in bar plots show mean+/-SEM.

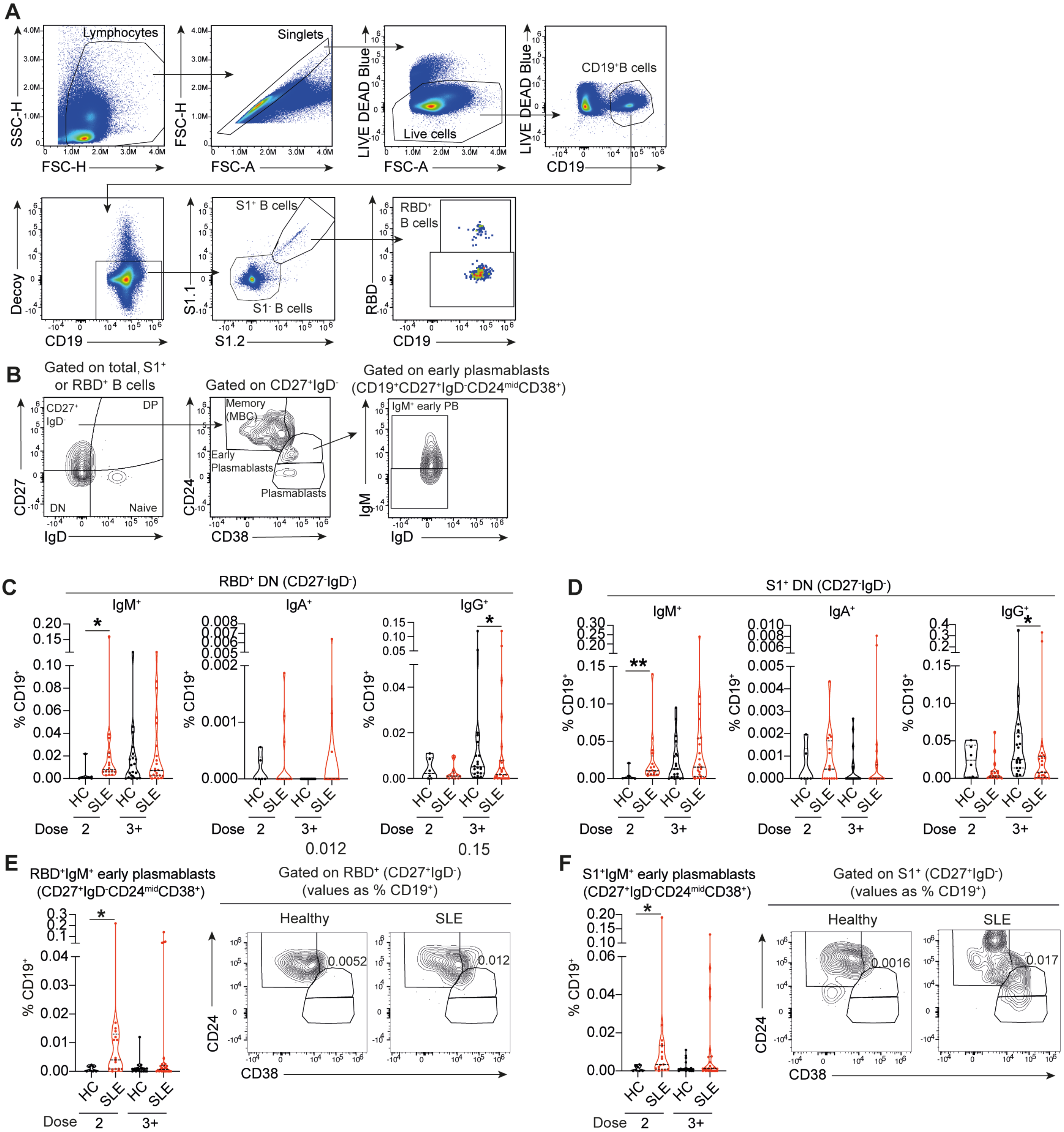


**Supplementary Figure 4.**

**(A)** Gating strategy for the identification of S1$^+$ and RBD$^+$ B cells. **(B)** Gating strategy for the identification of memory B cells (CD27$^+$IgD$^-$CD24$^+$CD38$^{lo}$) (MBC), IgM$^+$ early plasmablasts (CD27$^+$IgD$^-$CD24$^{mid}$CD38$^+$IgM$^+$), plasmablasts (CD27$^+$IgD$^-$CD24$^-$CD38$^+$), DP B cells (CD27$^-$IgD$^-$) and DN B cells (CD27$^-$IgD$^-$). **(C,D)** Cumulative data violin plots show the frequencies of (C) RBD$^+$ and (D) S1$^+$ double-negative (DN, CD27$^-$IgD$^-$) B cells within the CD19$^+$ population, in SLE patients and healthy controls (HC) after 2 or 3$^+$ doses of vaccine. **(E,F)** Representative contour plots and cumulative data violin plots showing the frequencies of (E) RBD$^+$ and (F) S1$^+$ IgM$^+$ early plasmablasts (CD27$^+$IgD$^-$CD24$^{mid}$CD38$^+$IgM$^+$) in SLE patients and healthy controls (HC) after 2 or 3$^+$ doses of vaccine Values in gates indicate the percentage within CD19$^+$ cells. **P*<0.05, ***P*<0.01, by unpaired t-test, Mann-Whitney test or data transformation according to data distribution and features (see methods). Violin plots display the median, along with the 25th and 75th percentiles.

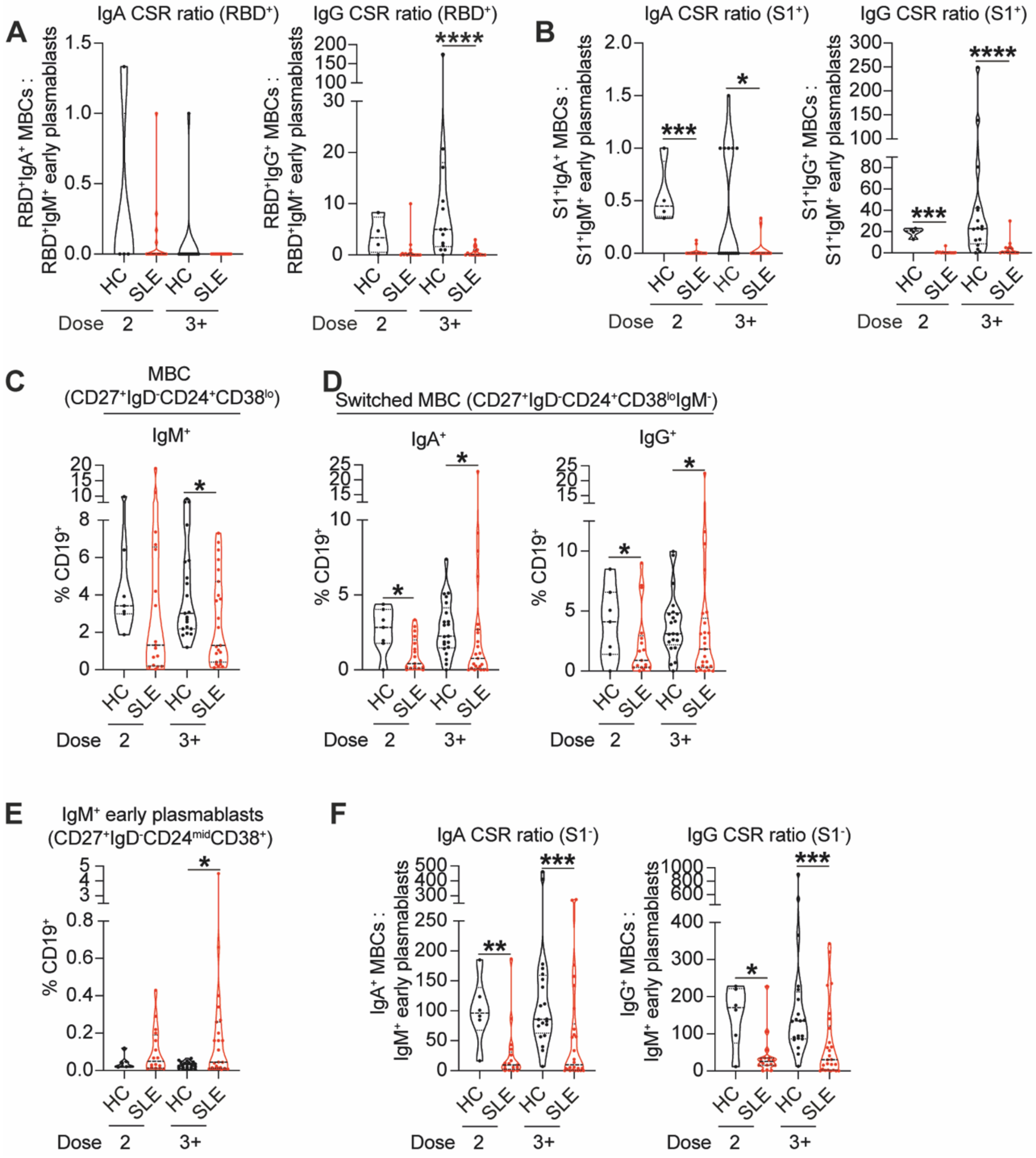


**Supplementary Figure 5.**

**(A,B)** Cumulative data violin plots showing the ratios of (A) RBD$^+$ IgA$^+$ and IgG$^+$ MBCs to RBD$^+$IgM$^+$ early plasmablasts, and (B) S1$^+$ IgA$^+$/$^+$ and IgG$^+$ MBCs to S1$^+$IgM$^+$ early plasmablasts, in SLE patients and healthy controls after 2 or 3$^+$ doses of vaccine. **(C,D)** Cumulative data violin plots showing the frequencies of (C) total IgM$^+$ unswitched memory B cells (MBC, CD27$^+$IgD$^-$CD24$^+$CD38$^{lo}$IgM$^+$) and (D) total IgG$^+$ and IgA$^+$ switched memory B cells (MBC, CD27$^+$IgD$^-$CD24$^+$CD38$^{lo}$IgM$^-$IgA/G$^+$) within the CD19$^+$ population in SLE patients and healthy controls (HC) after 2 or 3$^+$ doses of vaccine. **(E)** Cumulative data violin plot showing the frequencies of IgM$^+$ early plasmablasts (CD27$^+$IgD$^-$CD24$^{mid}$CD38$^+$IgM$^+$) in SLE patients and healthy controls (HC) after 2 or 3$^+$ doses of vaccine. **(F)** Cumulative data violin plots showing the ratios of total IgA$^+$/IgG$^+$ MBCs to total IgM$^+$ early plasmablasts, in SLE patients and healthy controls (HC) after 2 or 3$^+$ doses of vaccine. *$P$<0.05, **$P$<0.01 ***$P$<0.001, ****$P$<0.0001, by unpaired t-test, Mann-Whitney test or data transformation according to data distribution and features (see methods). Violin plots display the median, along with the 25th and 75th percentiles.

| | HC (n=22) | | SLE (n=18) | |
|---|---|---|---|---|
| **Dose #** | 2 (n=17) | 3-4 (n=5) | 2 (n=12) | 3-4 (n=6) |
| Age mean | 32.3 | 40.4 | 42.23809524 | 44.9 |
| Age range | 24-55 | 27-56 | 25-66 | 28-80 |
| Sex (%F/M) | 70.6/29.4 | 80/20 | 90.5/9.5 | 100/0 |
| *Ethnicity* | | | | |
| White | 29.4 | 80.0 | 41.7 | 50.0 |
| Afro-Caribbean | 0.0 | 0.0 | 33.3 | 16.7 |
| South/East Asian | 0.0 | 20.0 | 25.0 | 33.3 |
| Other | 0.0 | 20.0 | 0.0 | 0.0 |
| No data | 70.6 | 0.0 | 0.0 | 0.0 |
| **Treatment** | | | | |
| HCQ | - | - | 75.0 | 66.7 |
| Pred | - | - | 66.7 | 66.7 |
| MTX | - | - | 0.0 | 0.0 |
| MMF | - | - | 16.7 | 16.7 |
| Aza | - | - | 25.0 | 33.3 |
| Other | - | - | 0.0 | 0.0 |
| No medication | - | - | 16.7 | 16.7 |
| **Vaccine type** | | | | |
| mRNA | 94.1 | 80.0 | 50.0 | 83.3 |
| non-mRNA | 5.9 | 0.0 | 41.7 | 0.0 |
| Combination | 0.0 | 20.0 | 8.3 | 16.7 |
| No data | 0.0 | 0.0 | 0.0 | 0.0 |
| **Clinical features** | | | | |
| GS (range, average) | - | - | 0-14, 5.3 | 0-2, 1.0 |
| dsDNA (range, average, not determined) | - | - | <0.6-379, 56.6, 0 | 0.7-108, 25.6, 16.7 |
| C3 (range, average, not determined) | - | - | 0.7-1.2, 0.9, 0 | 0.6-1.3, 1.1, 0.0 |
| **BILAG (% A/B/C/D/E)** | | | | |
| General | - | - | 0/0/0/66.7/33.3 | 0/0/0/100/0 |
| Cutaneous | - | - | 8.3/8.3/0/75/8.3 | 0/0/0/83.3/16.7 |
| CNS | - | - | 8.3/0/0/41.7/50 | 0/0/0/66.7/33.3 |
| Musculoskeletal | - | - | 0/0/25/58.3/16.7 | 0/0/16.7/83.3/0 |
| Respiratory | - | - | 0/0/0/41.7/58.3 | 0/0/0/66.7/33.3 |
| GI | - | - | 0/0/0/33.3/66.7 | 0/0/0/66.7/33.3 |
| Ophthalmological | - | - | 0/0/0/25/75 | 0/0/0/50/50 |
| Renal | - | - | 8.3/8.3/0/66.7/16.7 | 0/0/50/33.3/16.7 |
| Hematological | - | - | 0/0/58.3/33.3/8.3 | 0/0/33.3/66.7/0 |

**Supplemental table 1. Demographics of longitudinal cohort of SLE patients and healthy controls**

HCQ = hydroxychloroquine, Pred = prednisolone, MTX = methotrexate, MMF = mycophenolate mofetil, Aza = azathioprine, GS = global score, dsDNA = double-stranded DNA, BILAG = British Isles Lupus Assessment Group, CNS = central nervous system, GI = gastrointestinal.

| | HC (n=65) | | SLE (n=59) | |
|---|---|---|---|---|
| Dose # | 2 (n=34) | 3-4 (n=31) | 2 (n=18) | 3-4 (n=34) |
| Age mean | 31.9 | 32.2 | 42.23809524 | 44.9 |
| Age range | 20-56 | 20-58 | 25-66 | 28-80 |
| Sex (%F/M) | 58.8/41.2 | 54.8/45.2 | 88.9/11.1 | 100/0 |
| **Ethnicity** | | | | |
| White | 47.1 | 74.2 | 38.9 | 52.9 |
| Afro-Caribbean | 0.0 | 0.0 | 38.9 | 5.9 |
| South/East Asian | 11.8 | 19.4 | 22.2 | 35.3 |
| Other | 5.9 | 6.5 | 0.0 | 5.9 |
| No data | 35.3 | 0.0 | 0.0 | 0.0 |
| **Treatment** | | | | |
| HCQ | - | - | 61.1 | 73.5 |
| Pred | - | - | 66.7 | 61.8 |
| MTX | - | - | 0.0 | 2.9 |
| MMF | - | - | 33.3 | 26.5 |
| Aza | - | - | 27.8 | 26.5 |
| Other | - | - | 0.0 | 0.0 |
| No medication | - | - | 16.7 | 11.8 |
| **Vaccine type** | | | | |
| mRNA | 91.2 | 83.9 | 72.2 | 55.9 |
| non-mRNA | 8.8 | 0.0 | 22.2 | 0.0 |
| Combination | 0.0 | 12.9 | 5.6 | 35.3 |
| No data | 0.0 | 3.2 | 0.0 | 8.8 |
| **Clinical features** | | | | |
| GS (range, average) | - | - | 0-17, 5.4 | 0-10, 1.8 |
| dsDNA antibody titres (range, average, not determined) | - | - | <0.6-397, 61.9, 0 | 0.7-326, 47.0, 14.7 |
| C3 (range, average, not determined) | - | - | 0.6-1.5, 0.9, 5.6% | 0.6-1.4, 1.0, 5.9% |
| **BILAG (% A/B/C/D/E)** | | | | |
| General | - | - | 0/0/0/77.8/22.2 | 0/0/0/94.1/5.9 |
| Cutaneous | - | - | 5.6/5.6/5.6/72.2/11.1 | 0/0/2.9/88.2/8.8 |
| CNS | - | - | 0/0/0/66.7/33.3 | 0/2.9/0/52.9/44.1 |
| Musculoskeletal | - | - | 0/11.1/11.1/66.7/11.1 | 0/0/14.7/85.3/0 |
| Respiratory | - | - | 0/0/0/55.6/44.4 | 0/0/0/64.7/35.3 |
| GI | - | - | 0/0/0/38.9/61.1 | 0/0/0/44.1/55.8 |
| Ophthalmological | - | - | 0/0/0/33.3/66.7 | 0/0/0/41.2/58.8 |
| Renal | - | - | 11.1/16.7/5.6/61.1/5.6 | 0/8.8/17.6/61.8/11.8 |
| Hematological | - | - | 0/0/50/44.4/5.6 | 0/0/47.1/52.9/0 |

**Supplemental table 2: demographics of SLE and healthy control cohorts used for SARS-CoV-2 blocking assays (MSD)**

HCQ = hydroxychloroquine, Pred = prednisolone, MTX = methotrexate, MMF = mycophenolate mofetil, Aza = azathioprine, GS = global score, dsDNA = double-stranded DNA, BILAG = British Isles Lupus Assessment Group, CNS = central nervous system, GI = gastrointestinal.

| | HC (n=28) | | SLE (n=40) | |
|---|---|---|---|---|
| Dose # | 2 (n=7) | 3-4 (n=21) | 2 (n=17) | 3-4 (n=23) |
| Age mean | 29.0 | 28.9 | 45.5 | 44.9 |
| Age range | 20-55 | 20-47 | 25-67 | 28-80 |
| Sex (%F/M/no data) | 42.9/57.1/0 | 52.4/42.9/4.7 | 88.2/11.8 | 100/0 |
| **Ethnicity** | | | | |
| White | 85.7 | 71.4 | 35.3 | 60.9 |
| Afro-Caribbean | 0.0 | 0.0 | 35.3 | 4.3 |
| South/East Asian | 0.0 | 19.0 | 23.5 | 21.7 |
| Other | 14.3 | 4.8 | 5.9 | 13.0 |
| No data | 0.0 | 4.8 | 0.0 | 0.0 |
| **Treatment** | | | | |
| HCQ | - | - | 52.9 | 69.6 |
| Pred | - | - | 70.6 | 60.9 |
| MTX | - | - | 0.0 | 8.7 |
| MMF | - | - | 29.4 | 21.7 |
| Aza | - | - | 23.5 | 21.7 |
| Other | - | - | 0.0 | 0.0 |
| No medication | - | - | 23.5 | 13.0 |
| **Vaccine type** | | | | |
| mRNA | 100.0 | 81.0 | 64.7 | 56.5 |
| non-mRNA | 0.0 | 0.0 | 29.4 | 4.3 |
| Combination | 0.0 | 14.3 | 0.0 | 39.1 |
| No data | 0.0 | 4.8 | 5.9 | 0.0 |
| **Clinical features** | | | | |
| GS (range, average) | - | - | 0-17, 5.8 | 0-10, 1.3 |
| dsDNA (range, average, not determined) | - | - | <0.6-397, 61.6, 0 | 0.7-326, 51.0, 13.0 |
| C3 (range, average, not determined) | - | - | 0.6-1.7, 1.0, 5.9 | 0.6-1.4, 1.0, 8.7 |
| **BILAG (% A/B/C/D/E)** | | | | |
| General | - | - | 0/0/0/76.8/23.5 | 0/0/0/95.7/4.3 |
| Cutaneous | - | - | 5.9/11.8/11.8/58.8/11.8 | 0/0/0/95.7/4.3 |
| CNS | - | - | 0/0/0/58.8/41.2 | 0/4.3/0/65.2/30.4 |
| Musculoskeletal | - | - | 0/11.8/11.8/64.7/11.8 | 0/0/17.4/82.6/0 |
| Respiratory | - | - | 0/0/0/64.7/35.3 | 0/0/0/69.6/30.4 |
| GI | - | - | 0/0/0/47.1/52.9 | 0/0/0/52.2/47.8 |
| Ophthalmological | - | - | 0/0/0/41.2/58.8 | 0/0/0/52.2/47.8 |
| Renal | - | - | 11.8/11.8/5.9/58.8/11.8 | 0/4.3/17.4/73.9/4.3 |
| Hematological | - | - | 0/0/52.9/41.18/5.9 | 0/0/30.4/60.9/8.7 |

**Supplemental table 3. demographics of SLE and healthy control cohorts used for *ex vivo* B cell phenotyping.**

HCQ = hydroxychloroquine, Pred = prednisolone, MTX = methotrexate, MMF = mycophenolate mofetil, Aza = azathioprine, GS = global score, dsDNA = double-stranded DNA, BILAG = British Isles Lupus Assessment Group, CNS = central nervous system, GI = gastrointestinal.

### SUPPLEMENTAL METHODS

#### Study population and BILAG global scoring system

Exclusion criteria for the study population were an age under 18, treatment with rituximab <2 year from the date of visit, participation in any interventional trial and pregnancy. Patients with severe central nervous system affectation, congestive heart failure, a history of cancer, severe glomerulonephritis, a history of recurrent or active infections such as HIV, tuberculosis, hepatitis B/C viruses were also excluded.

All participants underwent a structured examination by a rheumatologist to assess the organ disease involvement. Disease activity was assessed by the British Isles Lupus Assessment Group (BILAG), a standardized disease activity assessment [1]. Blood tests were performed as part of routine clinic visits and include: anti-dsDNA (double stranded DNA) autoantibody titres and complement C3 levels.

#### PBMC and serum isolation

Whole blood was collected in sodium heparin tubes (455051, Greiner Bio-One) and Serum SST tubes (456018, Greiner Bio-One). Non-coagulated whole blood was diluted 1:1 in PBS with 2% foetal bovine serum (FBS). Density gradient centrifugation using SepMate tubes (85450, STEMCELL Technologies) was used to isolate the PBMC fraction according to the manufacturer instructions. PBMCs were counted and viability estimated using a trypan blue exclusion assay. Cells were then resuspended at $10x10^6$ cells per ml in FBS containing 10% DMSO and stored in liquid nitrogen. Serum SST tubes were centrifuged at 1200g for 10 minutes, then serum was aliquoted into cryovials and stored at -80 ºC.

#### Quantification of wild type SARS-CoV-2 RBD-specific and S1-specific IgG, IgA and IgM titres

96-well Maxisorp plates (442404, ThermoFisher) were coated with 50 µl of carrier free-RBD (793604, Biolegend) or -S1 (792906, Biolegend) protein diluted to 2 µg/ml in PBS, sealed, and left overnight at 4°C. Plates were then washed 3 times with 250 µl of PBS with 0.05% Tween 20 (P1379, Sigma) (PBS-T). Plates were then blocked for at least 1 hour at room temperature with 250 µl 3% (w/v) skimmed milk powder (84615.0500, VWR) diluted in PBS-T.

For both RBD and S1, 100 µl of corresponding standard isotype, IgM (Ab01680-15-0, Absolute Biotech, clone CR3022), IgG (ab273073, Abcam, clone CR3022) and IgA (Ab01680-16-0, Absolute Biotech, clone CR3022), was added in duplicate. Standards concentrations ranged from 500 to 3.9 ng/ml for IgA and from 60 to 0.47 ng/ml for IgG and IgM. 100 µl of each pre-diluted sample, including a positive and a negative control, was also added to the plates in duplicate. The positive control was a known positive samples serum sample added to each plate in every experiment to determine intra-experimental variation; the negative control was a blank to which no serum sample was added, used for calculating background. For RBD-specific ELISAs, serum was diluted 1:50, 1:100, and 1:1000 for IgA, IgM, and IgG, respectively; for S1-specific ELISAs, serum was diluted 1:50, 1:50, 1:4000 for IgA, IgM, and IgG, respectively. Sample dilutions and standards concentration range were determined by titration experiments or from previous experiments. Samples and standards were diluted in 1% (w/v) skim milk in PBS-T. After standards, samples, and controls were added, plates were covered and left for two hours at room temperature.

Plates were then washed 3 times with 250 µl PBS-T. 50 µl of the corresponding anti-human HRP-conjugated secondary antibody, anti-human IgG (A18817, ThermoFisher), anti-human IgM (A18835, ThermoFisher), anti-human IgA (A0295-1ML, Sigma-Aldrich/Merk), diluted 1:3000 in 1% (w/v) skim milk powder in PBS-T, was then added. After residual bubbles were removed, plates were covered and left at room temperature for one hour. Plates were then washed 5 times with 250 µl PBS-T. 10 ml of o-phenylenediamine dihydrochloride (OPD) solution was prepared per plate by dissolving 1 tablet of OPD (11879250, FisherScientific) diluted in 9 ml of MilliQ $H_2O$ and then adding 1 ml stable peroxide buffer (11889270, FisherScientific). 100 µl of OPD was added to the wells of each plate. After no more than 10 minutes from the addition of OPD to the first wells, 50 µl of stop solution, 2N $H_2SO_4$, was added to each well. After residual bubbles were removed, the plates were then read in a plate reader (BioTek) with software BioTek Gen5 (v3.12) at an absorbance of 490 nm.

The average OD of the blank was subtracted from each sample and the duplicate sample ODs were averaged. The standard curve was interpolated using a Sigmoidal 4PL equation in Prism (GraphPad v9). This value was then multiplied by the dilution factor used for the chosen immunoglobulin isotype examined.

Upper and lower limits of values interpolated by the standard curve were determined by the linear phase of the curve. The upper and lower thresholds of the linear phase were determined, and samples with an OD above the upper threshold were assigned the concentration of the top standard. Samples with an OD below the lower threshold were assigned a concentration of 0 or of the concentration of lower threshold of the linear phase if they were at least 3x higher than the blank OD.

**Quantification of Omicron SARS-CoV-2 RBD-specific IgG and IgA titres**

An omicron strain B.1.1.529 RBD recombinant protein (SPD-C522e, AcroBiosystems) was used to coat the plates diluted to 2 µg/ml in PBS. An RBD-omicron-specific IgG monoclonal antibody against the B.1.1.529 strain (SPD-M415, AcroBiosystems) was used for the standard curve. Dilution of the serum for this assay was 1:2000. The rest of the protocols and reagents were as for the WT-RBD-IgG ELISA described above.

Serum RBD-omicron-specific IgA was determined using a kit (RAS-T099, AcroBiosystems) according to the manufacturer instructions using a 1:100 dilution.

**Total Ig ELISAs**

To measure the total isotype (IgG, IgA, and IgM) immunoglobulin concentration in serum samples, a sandwich ELISA (88-50550-22 for IgG, 88-50600-88 for IgA, 88-50620-88 for IgM, Invitrogen) was performed according to the manufacturer's instructions. A titration experiment was performed for each immunoglobulin isotype to determine the optimal dilution, which were ultimately determined to be 1:40,000 for IgA, 1:5,000 for IgM, and 1:50,000 for SLE patients and 1:500,000 for HC for IgG.

**Antigen-specific B cell phenotyping**

SARS-CoV-2 antigen-specific B cells were identified by tagging them with the subunit 1 (S1) of the spike protein (S) of the ancestral SARS-CoV-2 and the receptor-binding domain (RBD) which is contained within the S1. Biotin-conjugated S1 was coupled with two fluorochrome-labelled streptavidin to form two different S1-fluorochome conjugates. Biotin-S1 (793806, Biolegend) was conjugated with streptavidin-BV421 (405225, Biolegend) and streptavidin-APC (405207, Biolegend) separately, in PBS at a ratio of 1:6 (streptavidin-flurochrome:biotin-S1). Biotin-conjugated RBD (793904, Biolegend) was conjugated with streptavidin-BUV737 (612775, BD Biosciences) in PBS at ratio of 1:4. A decoy conjugate consisting of only biotin was constructed by mixing D-biotin and streptavidin-PE-Cy5 (405205, Biolegend) at a ratio of 1:40 (streptavidin-PE-Cy5:free biotin). These conjugates were incubated under agitation at 4°C for one hour.

5 x $10^6$ PBMCs were stained per sample. Cells were centrifuged at 500 g for 7 minutes at 4°C and washed once with PBS. Cells were incubated in 200 µl of LIVEDEAD™ fixable blue (L23105, Invitrogen) diluted 400-fold in PBS at 4°C in the dark for 30 minutes and FcR blocking agent (130-059-901, Miltenyi). Cells were then washed with FACS buffer (PBS 2% FBS and 0.2µM EDTA). 200 µl of 5µM D-biotin FACS buffer (D-biotin FACS buffer) containing 5ng of Biotin-PE-Cy5 was added and incubated for 30 minutes in the dark at 4°C. Following incubation with the decoy, cells were washed twice with D-biotin FACS buffer. Cells were then stained with the antigen probe cocktail (0.5µg/samples for both S1 and 0.25µg/sample for RBD) and surface antibodies (described in supplemental methods table 1) in 200 µl of D-biotin FACS buffer and left in the dark at 4°C for one hour to incubate. FACS tubes were agitated every 20 minutes during this incubation period to ensure maximal staining. Cells were then washed with D-biotin FACS buffer before 200 µl of IC Fixation Buffer (eBioscience) was added to each FACS tube and left to incubate for 30 minutes at 4°C in the dark. Following fixation, cells were washed twice with FACS buffer and resuspended in FACS buffer for acquisition. Samples were acquired in a CytekTM Aurora cytometer (5 lasers) using SpectroFlo (v3.0.3) with automated unmixing. Flow cytometric data were analysed using FlowJo software (v10.8.1).

| Manufacturer | Cat.no | Marker | Clone | Fluorochrome | Dilution |
|---|---|---|---|---|---|

| | | | | | |
|---|---|---|---|---|---|
| BioLegend | 302240 | CD19 | HIB19 | BV785 | 1:200 |
| BD Bioscience | 740291 | CD27 | M-T271 | BUV395 | 1:200 |
| BD Bioscience | 561315 | IgD | IA6-2 | PerCP-Cy5.5 | 1:200 |
| BioLegend | 356642 | CD38 | HB-7 | BV605 | 1:200 |
| BioLegend | 311120 | CD24 | ML5 | PE-Cy7 | 1:200 |
| BioLegend | 314546 | IgM | MHM-88 | APC-Fire 750 | 1:200 |
| Miltenyi | 130-116-882 | IgA | REA1014 | PE-vio615 | 1:200 |
| BD Bioscience | 555786 | IgG | G18-145 | FITC | 1:20 |

**Supplemental methods table 1: *Ex vivo* B cell phenotyping surface antibodies**

**IFNα CSR *in vitro* culture**

PBMCs from healthy controls were thawed, and total B cells were obtained by negative selection using the EasySep™ Human B Cell Isolation Kit (17954, StemCell Technologies). Total B cells were then stained for further flow cytometry sorting with anti-CD19 (302240, Biolegend), anti-CD27 (356430, Biolegend), anti-IgD (561315, BD). Viability staining using LIVE/DEAD™ Fixable Near IR Viability Kit (L34975, Invitrogen) was performed prior of the surface staining. Naive B cells ($CD27^-$ $IgD^+$) were sorted by flow cytometry using a BD FACSAria™ Fusion and BD FACSDiva (v9.4) and collected in recovery media (50% heat-inactivated FBS + 50% RPMI 1640 (R7388, Sigma-Aldrich)) at 4°C. Sorted naive B cells, 250k cells per well, were then cultured *in vitro* under IgG polarising class-switch consisting of complete RPMI (10% heat-inactivated foetal bovine serum and 1% penicillin/streptomycin), 1μg/ml anti-IgM (309-006-043, Jackson ImmunoResearch,), 1μg/ml CD40L (ALX-522-110-C010, Enzo Life Sciences) and 50ng/ml IFNγ (285-IF-100/CF, Bio-Techne) in a round bottom 96 well plate at 37°C and 5% $CO_2$ for 6 days. IgG polarising class-switch medium was refreshed on day 3 by gently centrifuging the cells (300g for 10min) and adding fresh media including the above-mentioned factors.

**Flow cytometry staining and analysis of *in vitro* cultures**

Cultured B cells in IgG polarising class-switch medium from each donor were collected after 6 days of incubation and stained for subsequent flow cytometry measurement. Cells were incubated in 200 μl of LIVEDEAD™ fixable blue (L23105, Invitrogen) diluted 400-fold in PBS at 4°C in the dark for 30 minutes and FcR blocking agent (130-059-901, Miltenyi). After washing in FACS buffer, cells were stained extracellularly with anti-CD19, anti-CD27, anti-CD24 and anti-CD38 (supplemental methods table 2) for 30 minutes in the dark at 4°C. Cells were then fixed and permeabilised using the eBioscience™ Intracellular Fixation & Permeabilization Buffer Set (88-8824-00, Invitrogen) and stained intracellularly with anti-IgD, anti-IgM, anti-IgG1, anti-IgG2, anti-IgG3 (supplemental methods table 3). Following intracellular staining, cells were washed twice with FACS buffer and resuspended in FACS buffer for acquisition. Samples were acquired in a CytekTM Aurora cytometer (5 lasers) using SpectroFlo software version 3.0.3 with automated unmixing. Analysis was performed using FlowJo version 10.8.1.

| Manufacturer | Cat.no | Marker | Clone | Fluorochrome | Dilution |
|---|---|---|---|---|---|
| BioLegend | 302240 | CD19 | HIB19 | BV785 | 1:200 |
| BD Bioscience | 740291 | CD27 | M-T271 | BUV395 | 1:200 |
| BioLegend | 356642 | CD38 | HB-7 | BV605 | 1:200 |
| BioLegend | 311120 | CD24 | ML5 | PE-Cy7 | 1:200 |

**Supplemental methods table 2: *In vitro* CSR culture surface marker antibodies**

| Manufacturer | Cat.no | Marker | Clone | Fluorochrome | Dilution |
|---|---|---|---|---|---|
| BD Bioscience | 561315 | IgD | IA6-2 | PerCP-Cy5.5 | 1:200 |
| Biolegend | 314546 | IgM | MHM-88 | APC-Fire 750 | 1:200 |
| Cytogonos | CYT-IGG1PE | IgG1 | SAG1 | PE | 1:200 |
| Cytogonos | CYT-IGG2PE | IgG2 | SAG2 | PE | 1:200 |
| Cytogonos | CYT-IGG2F | IgG2 | SAG2 | FITC | 1:200 |
| Cytogonos | CYT-IGG3F | IgG3 | SAG3 | FITC | 1:200 |

**Supplemental methods table 3: *In vitro* B cell CSR intracellular antibody table**

#### Patient and public involvement

SLE patients' organizations and a charity (LUPUS UK) were involved in the development of the main research question. SLE patients were highly interested in knowing the level of protection that SARS-CoV-2 vaccines provide, so they could return to their normal lives as soon and as safely as possible. At the time of consent, patients agreed to the burden of intervention and the time required to participate in the research which was only routine clinic visits. We plan to communicate the results of this study to SLE patients through both clinicians and patient meetings, aiming to inform them so they, together with their doctors, can make the best decisions regarding SARS-CoV-2 and other vaccinations, and possible future pandemics.

### REFERENCES:

1 Yee CS, Cresswell L, Farewell V, *et al*. Numerical scoring for the BILAG-2004 index. *Rheumatology*. 2010;49. doi: 10.1093/rheumatology/keq026
